# Corticosteroids for infectious critical illness: A multicenter target trial emulation stratified by predicted organ dysfunction trajectory

**DOI:** 10.1101/2024.03.07.24303926

**Authors:** Suraj Rajendran, Zhenxing Xu, Weishen Pan, Chengxi Zang, Ilias Siempos, Lisa Torres, Jie Xu, Jiang Bian, Edward J. Schenck, Fei Wang

**Affiliations:** Tri-Institutional Computational Biology & Medicine Program, Cornell University, NY, USA; Division of Health Informatics, Department of Population Health Sciences, Weill Cornell Medicine, New York, NY, USA; Division of Pulmonary and Critical Care Medicine, Department of Medicine, New York-Presbyterian Hospital-Weill Cornell Medical Center, Weill Cornell Medicine, New York, NY, USA; First Department of Critical Care Medicine and Pulmonary Services, Evangelismos Hospital, National and Kapodistrian University of Athens Medical School, Athens, Greece; Department of Health Outcomes and Biomedical Informatics. College of Medicine. University of Florida. Gainesville, FL, USA

## Abstract

Corticosteroids decrease the duration of organ dysfunction in a range of infectious critical illnesses, but their risk and benefit are not fully defined using this construct. This retrospective multicenter study aimed to evaluate the association between usage of corticosteroids and mortality of patients with infectious critical illness by emulating a target trial framework. The study employed a novel stratification method with predictive machine learning (ML) subphenotyping based on organ dysfunction trajectory. Our analysis revealed that corticosteroids’ effectiveness varied depending on the stratification method. The ML-based approach identified four distinct subphenotypes, two of which had a large enough sample size in our patient cohorts for further evaluation: “Rapidly Improving” (RI) and “Rapidly Worsening,” (RW) which showed divergent responses to corticosteroid treatment. Specifically, the RW group either benefited or were not harmed from corticosteroids, whereas the RI group appeared to derive harm. In the development cohort, which comprised of a combination of patients from the eICU and MIMIC-IV datasets, hazard ratio estimates for the primary outcome, 28-day mortality, in the RW group was 1.05 (95% CI: 0.96 - 1.04) whereas for the RW group, it was 1.40 (95% CI: 1.28 - 1.54). For the validation cohort, which comprised of patients from the Critical carE Database for Advanced Research, estimates for 28-day mortality for the RW and RI groups were 1.24 (95% CI: 1.05 - 1.46) and 1.34 (95% CI: 1.14 - 1.59), respectively. For secondary outcomes, the RW group had a shorter time to ICU discharge and time to cessation of mechanical ventilation with corticosteroid treatment, where the RI group again demonstrated harm. The findings support matching treatment strategies to empirically observed pathobiology and offer a more nuanced understanding of corticosteroid utility. Our results have implications for the design and interpretation of both observational studies and randomized controlled trials (RCTs), suggesting the need for stratification methods that account for the differential response to standard of care.

## Introduction

Infection is the most common cause of critical illness across the globe. Critically ill patients with infection present with organ dysfunction that manifest in overlapping and summative phenomena including circulatory, kidney and lung failure^1^. These organ specific failures are subject to their own syndromic definitions, such as septic shock in the case of circulatory failure and the acute respiratory distress syndrome (ARDS) in the case of hypoxemic respiratory failure. However, patients with septic shock often meet the syndromic definition of ARDS and vice versa. Both septic shock and ARDS have been evaluated in numerous epidemiologic and clinical studies with a marked lack of breakthrough therapies^2–4^. Moreover, the organ specific source of infection, e.g. pneumonia, have their own distinct evidentiary base. To reconcile these conflicting conceptual models of the causes of and consequences of infectious critical illness, there has been a call to redefine critical illnesses as phenotypic representations of treatable traits^1^.

Corticosteroids are an adjunctive therapy for selected patients with infectious critical illness due to their vasoconstrictive, anti-inflammatory, and immunomodulatory effects^5,6^. These effects lead to an empirical reduction in the duration of organ dysfunction in septic shock, and ARDS^7^. However, the benefits of corticosteroid treatment in infectious critical illness are to be balanced with immunosuppression, hyperglycemia, and other adverse events^8–10^. There remains an unmet need to establish a more precise target population within infectious critical illness to improve the risk benefit profile of corticosteroid therapy^7,11–13^.

Given the landscape of overlapping syndromes and causes of infectious critical illness, the development of a precision medicine framework is needed^1^. Recent attempts to evaluate heterogeneity of corticosteroid treatment effects have used specific syndromes, e.g. septic shock^14,15^, or specific infectious causes of critical illness. e.g. COVID-19^16,17^. However, these analyses are restricted to specific syndromes and do not explicitly explore the corticosteroids on expected duration of organ dysfunction. Moreover, the narrow analyses do not reflect the broad population of infectious related critical illness treated with steroids in clinical practice^18^. To bridge this gap, there is a need to evaluate the effect of corticosteroids as used in clinical practice using target trial emulation^19,20^. More specifically, since the observed beneficial effect of corticosteroids in infectious critical illness is through the amelioration of organ dysfunction, defining treatment heterogeneity between subphenotypes of predicted organ dysfunction trajectory may have value^21^.

We conducted a retrospective multicenter study to explore the effects of corticosteroids in patients with infectious critical illness, through a target trial emulation framework stratified by subphenotypes of predicted organ dysfunction trajectory (**Figure 1**). Specifically, the objectives of this study were to (1) determine whether the effectiveness of corticosteroid administration on 28-day mortality is modified by stratification based on trajectory-based subphenotypes; and (2) evaluate the effectiveness of corticosteroids on secondary endpoints including the duration of ICU stay and the duration of mechanical ventilation. This real-world data will serve as a model to demonstrate the feasibility of a subphenotype stratified clinical trial based on a simple mechanistic rationale that takes the expected response to standard of care into account.

**Figure 1.**
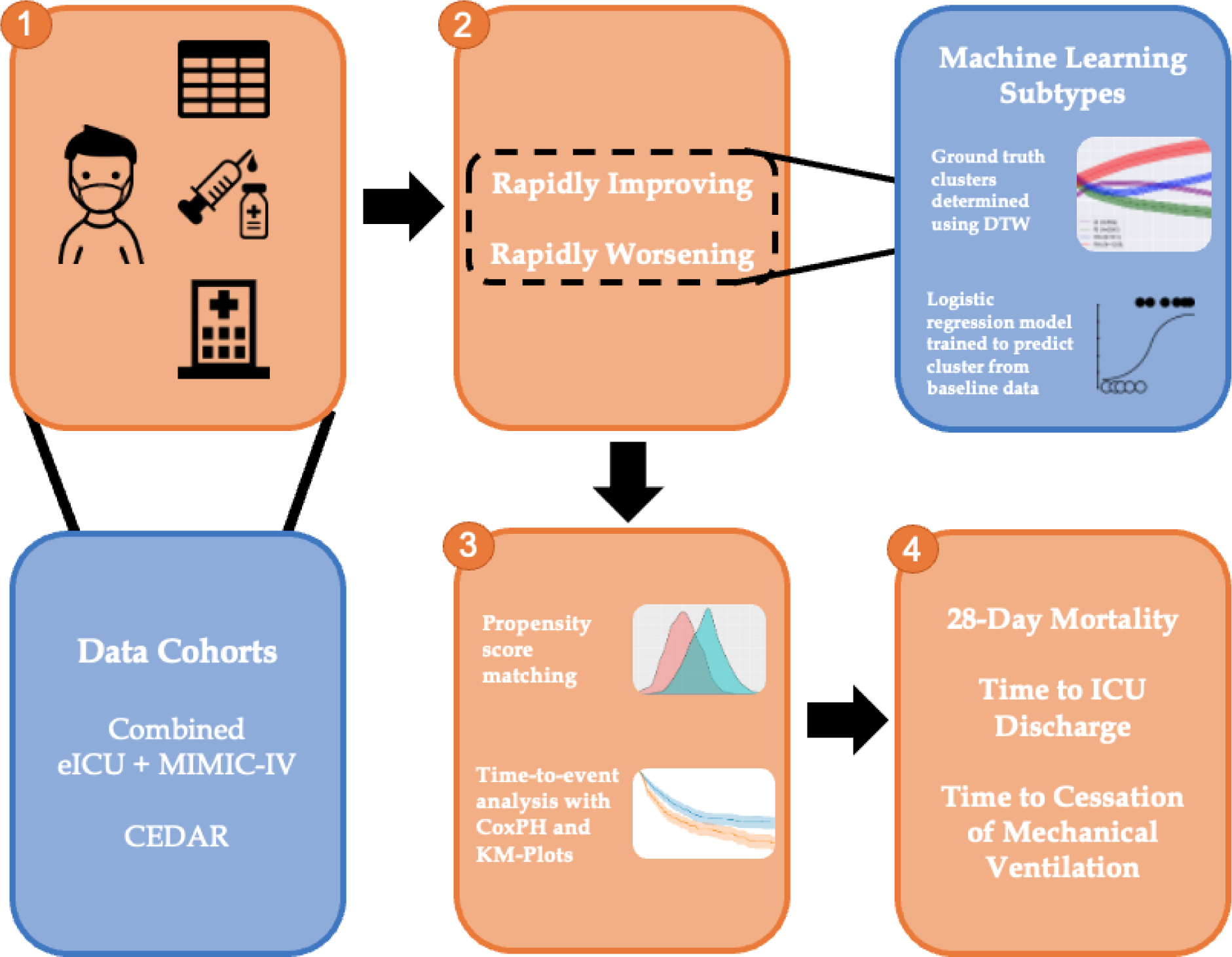
Overview of analyses conducted in study. Relevant covariates and patient information were extracted from multiple data cohorts. These patients were then stratified and processed as per trial emulation setup. Statistical methods were then used to determine relevant outcomes.

## Methods

### Data Source and Cohort

Three distinct data sources including Medical Information Mart for Intensive Care (MIMIC-IV)^22^, eICU^23^, and Critical carE Database for Advanced Research (CEDAR)^24^ were used for our analysis. The MIMIC-IV database was derived from Beth Israel Deaconess Medical Center (BIDMC), which is a teaching hospital of Harvard Medical School in Boston, Massachusetts with 673 licensed beds, including 493 medical/surgical beds, 77 critical care beds, and 62 OB/GYN beds. This database covered a decade of admissions between 2008 and 2019. The eICU database was built based on multi-center data from patients who were admitted to one of 335 units at 208 hospitals located throughout the US between 2014 and 2015. Both MIMIC-IV and eICU databases are publicly available data sources. The CEDAR database was built on New York-Presbyterian/Weill Cornell Medical Center (NYP/WCMC), which included 862 beds in total. This database is private and includes ICU admissions dating from 2001-2020. We used the public datasets integrating the MIMIC-IV and eICU as a development cohort to perform our analysis and validated them on the private database (CEDAR).

### Target Trial Emulation and Specification

The study employs an intention-to-treat (ITT) analysis. We defined infectious critical illness through modification of the Sepsis-3 criteria^25^. We maintained the need for a suspected infection (combination of administration of antibiotics and a body fluid culture specimen obtained). We established any total SOFA score of at least 2, instead of an increase of 2 over a baseline, as having an infectious critical illness. The enrollment window was defined as the 24-hour period after ICU admission. The eligibility criteria for inclusion in the study were as follows: patients aged 18 or older at enrollment window (baseline), a diagnosis of sepsis at enrollment window as defined by our infectious critical illness criteria, no history of suspected infection before the enrollment window, and no prescription for corticosteroids before the enrollment window. The inclusion-exclusion cascade for the patients in our study is shown in **Supplemental Figures 1, 2, and 3**. Two treatment strategies were compared in this study. Strategy 0 involved no initiation of any corticosteroids drug at the enrollment window, while Strategy 1 involved the initiation of hydrocortisone at a dose of at least 160 mg per day at the enrollment window (comparable to 40 mg prednisone or 32 mg methylprednisolone), within a window of 10 hours before to 24 hours after ICU admission. We compute cumulative milligram dosing of hydrocortisone at the enrollment window, and if a patient received at least 160 mg per day hydrocortisone equivalent, they are denoted as having corticosteroid exposure. We consider corticosteroids including Prednisolone, Prednisone, Hydrocortisone, Dexamethasone, and Methylprednisolone. A table showing the breakdown of corticosteroid usage for treated patients is shown in **Supplemental Table 1**. In accordance with ITT analyses, patients who received 160 mg per day after the enrollment window were not censored and stayed assigned to their initial treatment group. The study emulated randomization by classifying individuals according to the strategy that their data were compatible with at baseline and adjusting for baseline confounders. We handled immortal time bias by doing the following: (1) Time zero for treated patients was set to time of treatment to remove immortal time between eligibility and exposure; (2) cloning all patients who died at the enrollment window (on Day 1) to both treatment strategies and conducted sensitivity analyses with their exclusion^26^.

Multiple types of baseline covariates including vital signs, laboratory measurements, and demographics were selected. The vital signs included heart rate, mean arterial pressure (MAP), respiratory rate, oxygen saturation (SpO2), systolic arterial blood pressure (Systolic ABP), and temperature. These variables were selected as they provide crucial information about the patient’s physiological status and are routinely monitored in ICU settings. Laboratory measurements comprised a broad range of biochemical, hematological, and physiological parameters. Biochemical measurements included albumin, alanine aminotransferase (ALT), aspartate aminotransferase (AST), bilirubin, blood urea nitrogen (BUN), chloride, creatinine, C-reactive protein (CRP), glucose, lactate, and sodium. Hematological measurements included bands, hemoglobin, international normalized ratio (INR), platelet count, and white blood cell (WBC) count. Physiological parameters included the fraction of inspired oxygen (FiO2), Glasgow Coma Scale (GCS) score, arterial oxygen partial pressure (PaO2), and urine output. These laboratory measurements were selected as they provide a comprehensive overview of the patient’s organ function and metabolic status. The SOFA score subcomponents, including respiration score, cardiovascular score, central nervous system (CNS) score, liver score, coagulation score, and renal score, were also included. These scores provide a quantifiable measure of organ dysfunction, vasopressor administration, and mechanical ventilation, which are key aspects of sepsis. Demographic data included age, sex, and body mass index (BMI). Age and BMI were quantized to facilitate analysis, with BMI categories defined according to the World Health Organization guidelines. In addition, The Elixhauser Comorbidity Index was used to evaluate comorbidities based on past medical history^27^. Regarding preprocessing those covariates, we removed some abnormal values if they were above the 99th percentile and used the median value to fill in missing values. Missingness of covariates are shown in **Supplemental Table 2**. If there are multiple values within the 24-hour enrollment window, the worst clinical condition values were selected.

The study examined one primary outcome: 28-day mortality from the time of randomization. Two secondary outcomes were also studied: time to ICU discharge from the time of randomization, and time to cessation of mechanical ventilation from the time of randomization. Time to mechanical ventilation cessation was defined as a period of 24 hours without either invasive or non-invasive ventilation support. For secondary outcomes, death was considered as a competing risk^28^. Each patient was followed from his/her baseline until the day of his/her death, loss to follow-up, or discharge, whichever occurred first (**Supplemental Figure 4**). The study aimed to estimate the observational analog of the intention-to-treat effect, providing a robust analysis of the potential impact of corticosteroid treatment on sepsis outcomes. More details about target trial emulation specifications are shown in **Supplemental Tables 3, 4, and 5**.

### Statistical Methods

To balance the distribution of baseline covariates between the treatment and control groups, the study employed a propensity score matching technique^29^. Propensity scores were estimated using a logistic regression model, with the treatment assignment as the dependent variable and the baseline covariates as independent variables. The propensity score for each patient, representing the probability of receiving the treatment given their baseline characteristics, was then added to the dataset for subsequent matching. The study implemented a nearest-neighbor matching algorithm based on the propensity scores, using a 1:4 matching ratio with repetition^30^. This approach allowed each treated patient to be matched with up to four control patients. The Mahalanobis distance metric was used to measure the similarity in propensity scores between treated and control patients. A caliper value, set as the median absolute deviation of the propensity scores, was used to define the maximum allowable difference in propensity scores for a match. This caliper was adjusted as necessary during the iterative matching process to ensure optimal balance in the covariates between the treatment and control groups. The process was repeated until all covariates (or all but 2% * number of covariates) were adequately balanced, as indicated by a standard mean difference (SMD) below 0.1^31^. This threshold is a common standard in observational studies, where an SMD of less than 0.1 is considered indicative of a negligible difference in the mean or prevalence of a covariate between the treatment groups.

The study evaluated the cumulative incidence of the different outcomes (28-day mortality, time to ICU discharge, and time to cessation of mechanical ventilation) in the treatment and control groups using a Cox proportional hazards model. This model allowed for the estimation of the hazard ratio (HR) between the treatment and control groups, providing a measure of the effect of the treatment on the time until the occurrence of the event of interest^32^. To account for competing events, the study adjusted the duration for different outcomes^28^. In the analysis of 28-day mortality, patients who were discharged before the 28-day mark had their time to event set to the maximum duration of 28 days. This adjustment assumes the best-case scenario for these patients, that they would have survived if they had stayed in the ICU for the full 28 days. Conversely, for the outcomes of time to ICU discharge and time to cessation of mechanical ventilation, patients who died before the 28-day mark had their duration set to 28 days. This adjustment assumes the worst-case scenario for these patients, that they would not have been discharged or had their ventilation ceased if they had survived to the end of the 28-day period. Based on this experimental setup, a HR of less than 1 is associated with benefits from steroids on all outcomes. 95% confidence intervals were estimated for each HR measurement.

### Cohort Specific Analyses

To address the characteristics of complexity and heterogeneity of sepsis, we stratified patients into predictive subgroups based on different strategies to investigate the differential effectiveness of corticosteroid treatment. We first identified patients as the RI and RW based on the previous Xu et al. study^21^, which defined sepsis patients using data within the first 6 hours of ICU admission, represented each patient with a vector of 12 SOFA scores obtained every 6 hours within the first 72 hours of ICU admission, and performed the DTW and hierarchical clustering to identify four subphenotypes including RI, RW, DI, DW. We used data from the entire 24-hour enrollment window to predict trajectory assignment. Once we obtained subphenotypes, logistic regression (LR) models were trained on these ground truth subphenotype labels with the predictors being baseline measurements and demographics of the patient. All variables used in training the model are shown in **Supplemental Text 1**. Both L1 and L2 penalties were applied to the LR with a mix ratio of 0.5. Class weights were used to ensure that the models did not overfit to the majority class. The models were trained on the development cohort and validated on the validation cohort to ensure generalizability. The trained LR model can be used to predict whether patients belong to RI and RW groups based on their baseline covariates. We focused on the RI and RW patients to define differential effects on sepsis subphenotypes with predicted divergent organ dysfunction.

### Sensitivity Analyses

To ensure the robustness of our results, we performed the following sensitivity analyses. (1) Strategy 1 is changed to initiation of hydrocortisone at a dose of 200 mg per day at enrollment window; (2) Steroid exposure period is limited to 0 - 24 hrs post ICU admission; (3) Exclusion of patients who die during the exposure period (Day 1) instead of cloning; (4) Excluding laboratory values with high percentage of untested patients (ALT, AST, Albumin, Bands, Troponin T, CRP); (5) Inclusion of source of infection as a covariate for 28-day mortality. (5) could only be performed on the eICU dataset as date of diagnoses are not provided by other cohorts and hence, we could not confirm whether diagnoses were given at baseline. Lastly, we perform (6), an analysis where we include patients that the clinician intended on treating during the enrollment period. By doing so, we include many patients who were primarily treated with hydrocortisone that received an initial dosage during the enrollment period but did not complete their full regiment till after the enrollment period. This change only added patients in the validation cohort (CEDAR) and was primarily patients who were treated with hydrocortisone.

## Results

### ML-Based Patient Stratification

Based on our previous study regarding the identification of sepsis subphenotypes with SOFA trajectory, we obtained four subphenotypes including Rapidly Worsening (RW), Delayed Worsening (DW), Rapidly Improving (RI) and Delayed Improving (DI) in our development cohorts (eICU and MIMIC-IV) and validation cohort (CEDAR). The trajectory characteristics of the subphenotype were shown in **Supplemental Figure 5**. In this study, to stratify patients into different groups and investigate the effects of corticosteroids, we selected RI and RW, which account for most of the patients. In the eICU-MIMIC dataset, we identified 5,038 RW patients and 11,239 RI patients. In the CEDAR dataset, we identified 1,368 RW and 2,843 RI patients. The RI patients were characterized by a decreasing SOFA score over time, while the RW patients displayed an increasing SOFA score over the same duration. Further, we built two logistic regression models for the RI and RW patients, respectively. Performance evaluations for the logistic regression can be found in **Supplemental Text 1**.

### Characteristics of Development and Validation Cohorts

Demographic details of the development and validation cohorts can be found in **Supplemental Table 6** and **7** respectively. The epidemiological profile of sepsis patients in the development cohort treated with steroids significantly differs from those who were untreated. In the treated cohort, patients tended to be slightly younger with a median age of 66 years, as opposed to 67 years in the untreated group, though this difference was non-significant. There was a more pronounced disparity in the comorbidity burden, as indicated by the Elixhauser index, with the treated group showcasing a higher median value of 6.0 compared to the untreated group’s median of 5.0 (p < 0.001). This suggests that the patients who received steroids possibly had a more complex clinical profile. Both groups had comparable length of stays in the ICU as well as baseline SOFA scores. When examining the sources of infections, both groups predominantly suffered from septicemia bacteremia, with a higher proportion seen in the untreated group (45.8%) relative to the treated (43.5%) (p < 0.001). Pneumonia was another significant source, affecting a larger fraction of the treated cohort (46.4%) compared to 28.8% in the untreated population (p < 0.001). Other sources of infection, including infections of the central nervous system, intra-abdominal region, skin soft tissue, and urinary tract, displayed varying proportions.

In the validation cohort, the age of the treated cohort stood out, with these patients being notably younger, having a median age of 64.0 years, in stark contrast to the untreated group’s median age of 72.0 years—a difference that was statistically significant (p < 0.001). Regarding sex, a slightly higher percentage of males were untreated (56.6%) compared to those treated (50.7%). The comorbidity burden was uniformly distributed with a median of 15.0 in both cohorts, suggesting similar underlying health complexities in both groups. Interestingly, the length of ICU stay was slightly shorter for the treated group, with a median of 6.0 days, compared to the untreated 7.2 days, though this did not reach statistical significance (p = 0.163). In terms of clinical severity, the mechanical ventilation rates at admission were similar across the groups, and the SOFA scores remained comparable, indicating a similar severity of illness upon admission. In terms of the sources of infections, the distribution was consistent between the treated and untreated groups. Pneumonia and septicemia bacteremia were dominant in both cohorts, with no significant differences in proportions.

The basic characteristics of development and validation cohorts are summarized in **Supplemental Table 8** in terms of three outcomes including 28-day mortality, time to ICU discharge, time to cessation of mechanical ventilation after balancing. For the outcome of 28-day mortality, we examined the 28-day mortality incidence and overall duration from ICU admission to death across various stratifications in both the development and validation cohorts. In the development cohort, the overall mortality incidence was approximately 17.7%, with an average duration from ICU admission to death of about 22.13 days. We also found that the RI subgroup displayed a lower mortality rate compared to the RW group in both the development and validation cohorts (**Supplemental Table 9**). For the outcome of time to ICU discharge, the overall discharge incidence was approximately 71.1% in the development cohort, with patients staying in the ICU for an average duration of about 11.83 days prior to discharge. In the development cohort, the RI group showed a higher discharge incidence compared to the RW group. In the validation cohort, the discharge incidence between the RI and RW group are comparable. For time to cessation of mechanical ventilation, the overall incidence of ventilation cessation was approximately 79.1%, with an average duration of about 8.70 days from ICU admission to the cessation of ventilation. In the development cohort, the incidence of ventilation cessation is comparable between RI and RW groups.

### Survival Analysis on Outcomes

After obtaining balanced emulated trials with propensity score matching technique, we computed the hazard ratio for the different outcomes across the various cohorts and stratifications. The SMD of covariates before and after balancing are depicted in **Supplemental Figure 6, 7, and 8**. **Supplemental Table 10** and **11** show the median and IQR values of all covariates before and after balancing for treated and untreated population. Note that, the cessation of ventilation outcome in the validation cohort was excluded from further trial emulation analyses due to the inadequate confounder balance. Kaplan Meier plots for all cohorts and outcomes in terms of RI and RW are shown in **Figure 2** and **Supplemental Figure 9**.

**Figure 2.**
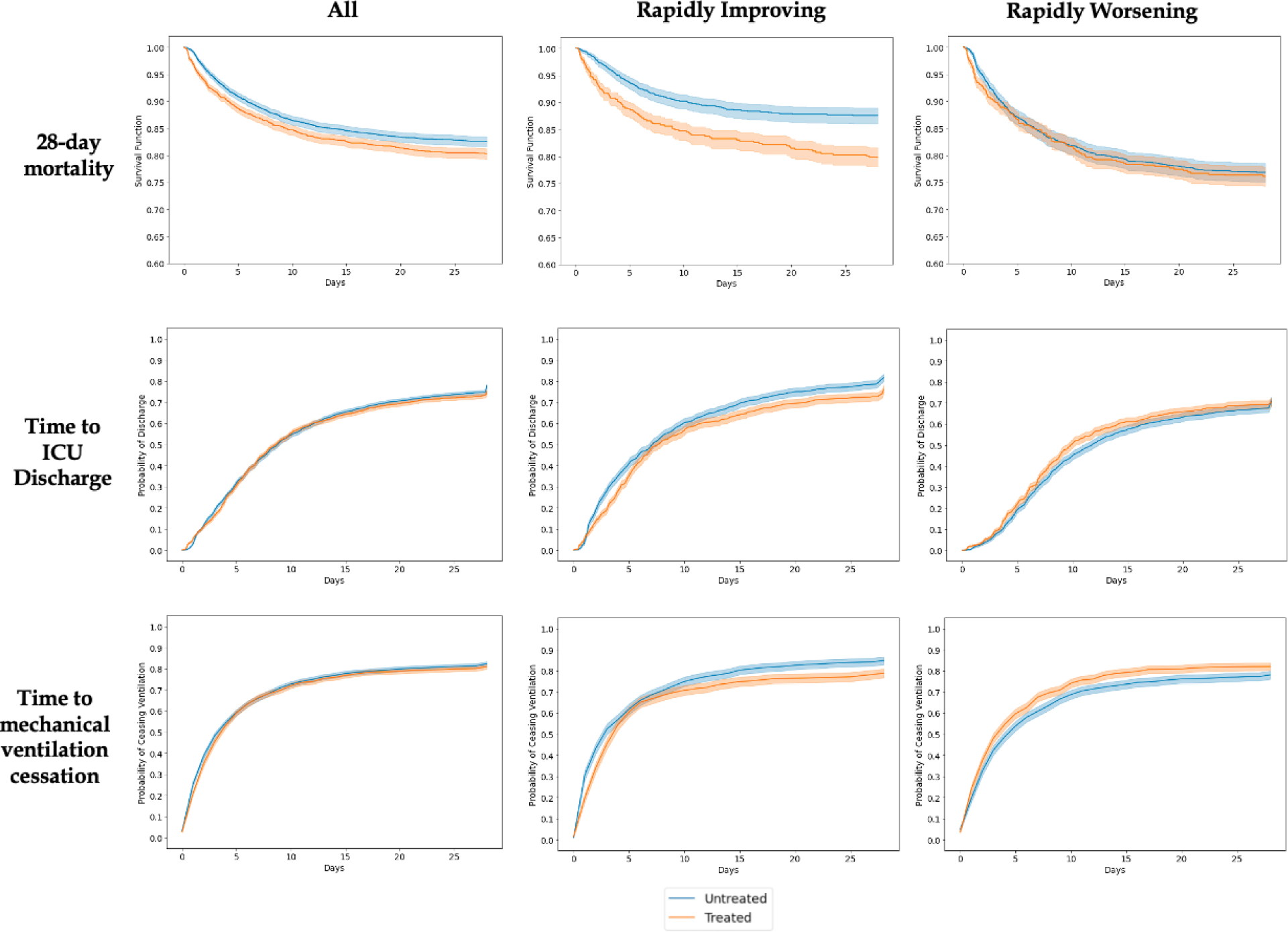
Kaplan-Meier plots for eICU-MIMIC cohorts. Shows all, rapidly improving, and rapidly worsening stratifications across different outcomes for both treated and untreated patients.

#### 28-Day Mortality

In the evaluation of 28-day mortality, the HR serves as an indicator of the relative risk of mortality within 28 days for patients treated with steroids compared to those who were not (**Figure 3**). An HR of less than 1 implies a decrease in 28-day mortality associated with steroid treatment. For the development cohort, the overall HR was 1.10 (95% CI: 1.04 - 1.16), suggesting a negative impact of steroids on 28-day mortality. The machine learning subgroups displayed pronounced differences. The Rapidly Improving group had an HR of 1.40 (95% CI: 1.28 - 1.54), suggesting an increased risk of mortality with steroid treatment. In contrast, the Rapidly Worsening group showed an HR of 1.05 (95% CI: 0.96 - 1.04). In th validation cohort, the hazard ratios were generally higher across all stratifications compared to the development cohort, indicating a potential increased risk of 28-day mortality associated with steroid use (**Supplemental Figure 10**). The overall hazard ratio (HR) was 1.48 (95% CI: 1.34 - 1.63), suggesting an overall detrimental effect of steroids on 28-day mortality. In the Rapidly Improving group, the HR was 1.34 (95% CI: 1.14 - 1.59), consistent with the increased mortality risk observed in other subgroups. The detrimental effect was attenuated in the Rapidly Worsening group, the HR was 1.24 (95% CI: 1.05 - 1.46), the lowest among all the stratifications in the validation cohort.

**Figure 3.**
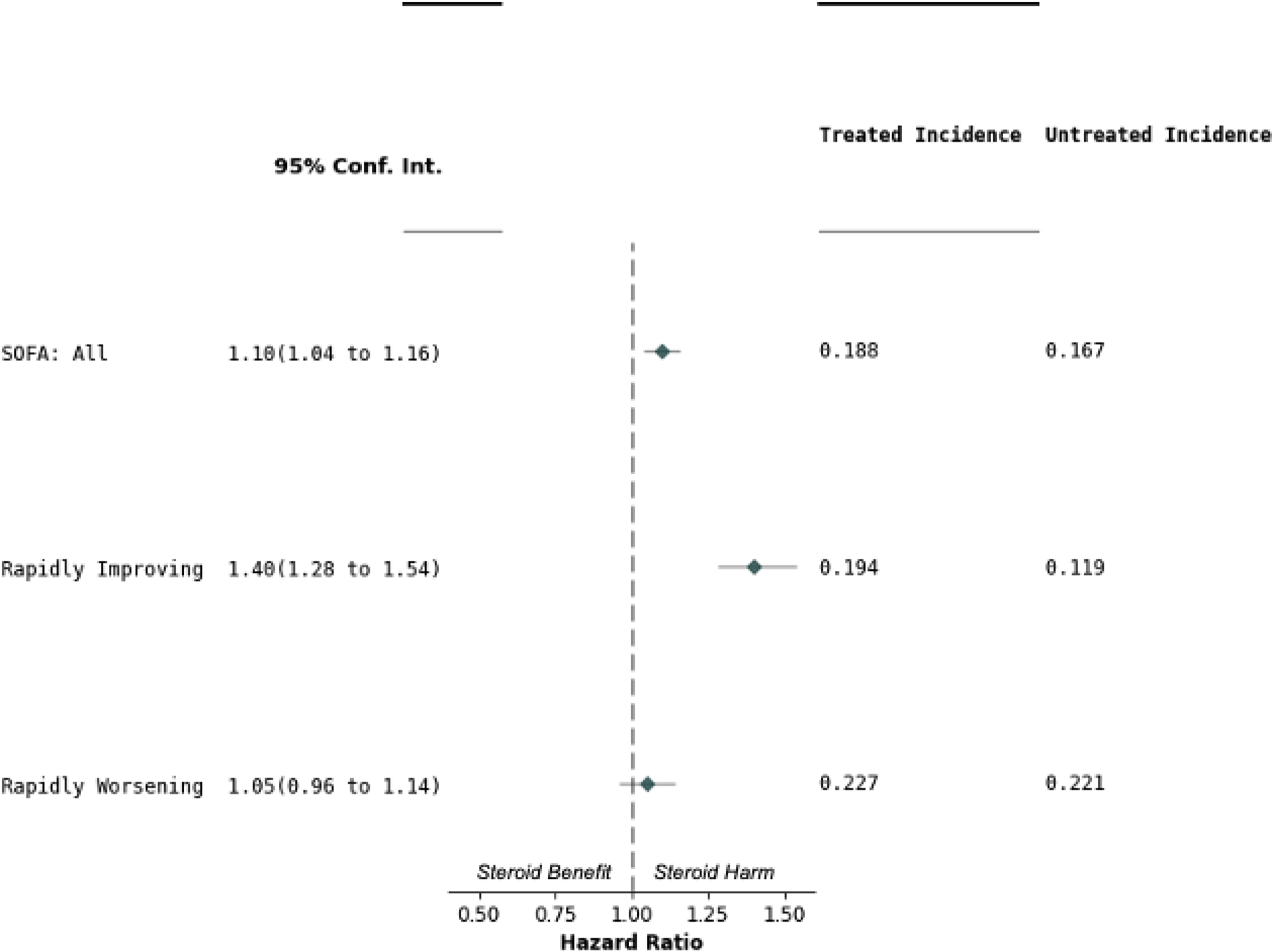
Hazard ratios and cumulative incidence for 28-day mortality outcome in eICU-MIMIC cohort. Forest plot shows the hazard ratios and 95% confidence intervals for each patient stratification. Dashed vertical line represents a hazard ratio of 0. Cumulative incidences for treated and untreated populations are shown on the right.

#### Time to ICU Discharge

In the evaluation of time to ICU discharge, a HR less than 1 suggests a beneficial effect of steroid treatment, as it indicates a shorter time to discharge for treated patients compared to those untreated (**Figure 4**). In the development cohort, the overall HR was 1.02 (95% CI: 0.98 - 1.07), suggesting a slightly prolonged time to discharge associated with steroid use. The machine learning subgroups showed divergent trends, with the Rapidly Improving group displaying an HR of 1.10 (95% CI: 1.01 - 1.18) and the Rapidly Worsening group showing an HR of 0.90 (95% CI: 0.83 - 0.97). In the validation cohort, th overall HR was 1.17 (95% CI: 1.09 - 1.25), indicating a slightly prolonged time to discharge with steroid use (**Supplemental Figure 11**). The Rapidly Improving groups had an HR significantly greater than 1, suggesting harm from steroids. Notably, the Rapidly Worsening group showed an HR of 1.00 (95% CI: 0.90 - 1.11), indicating no significant harm from steroid use.

**Figure 4.**
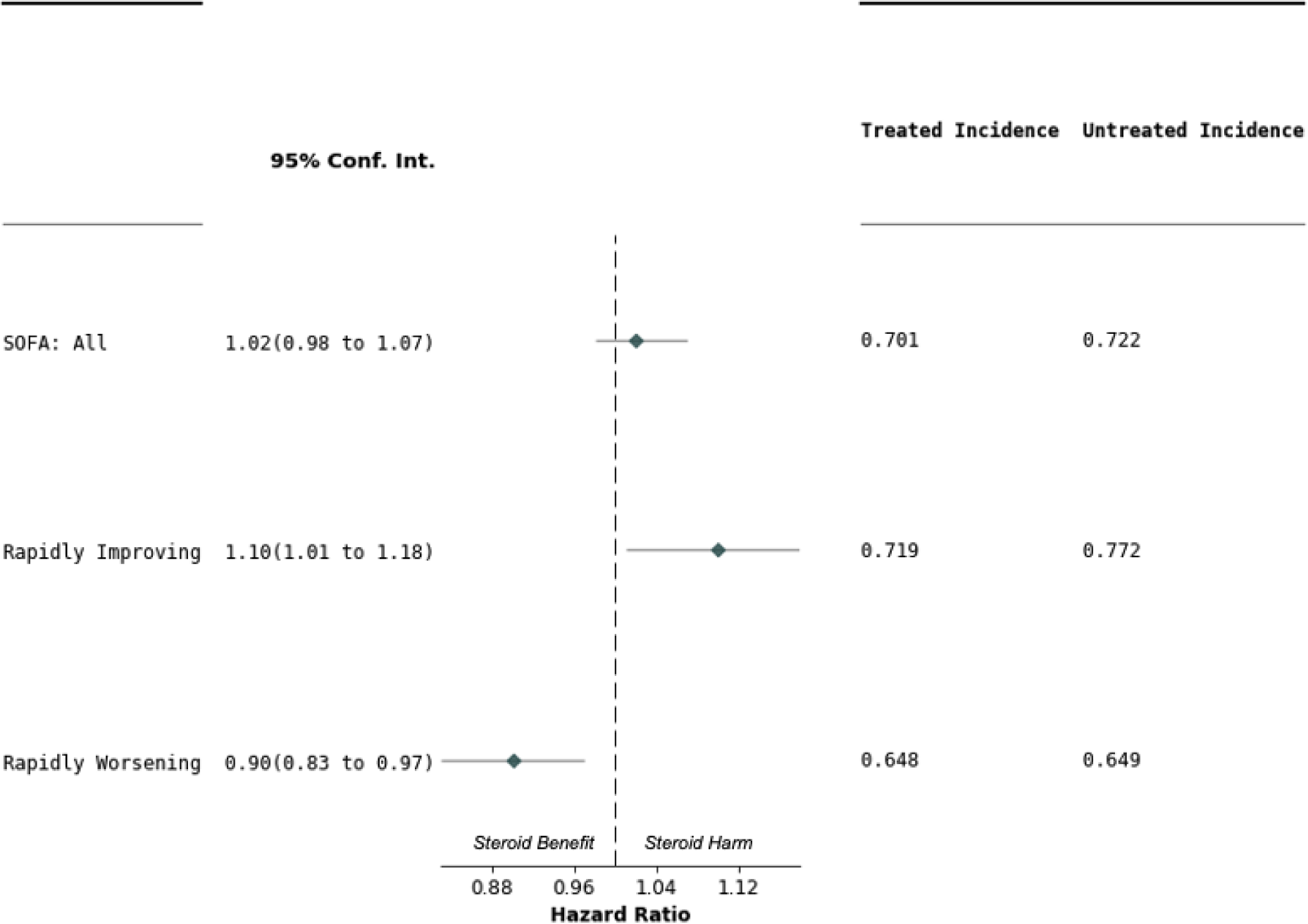
Hazard ratios and cumulative incidence for time to ICU discharge outcome in eICU-MIMIC cohort. Forest plot shows the hazard ratios and 95% confidence intervals for each patient stratification. Dashed vertical line represents a hazard ratio of 0. Cumulative incidences for treated and untreated populations are shown on the right.

#### Time to Cessation of Mechanical Ventilation

In assessing the time to cessation of mechanical ventilation, a HR less than 1 would indicate a beneficial effect of steroid treatment, as it would signify a shorter duration of ventilation for treated patients compared to those untreated (**Figure 5**). For the development cohort, the overall HR was 1.02 (95% CI: 0.98 - 1.07), suggesting no significant difference in ventilation cessation time associated with steroid use. The machine learning subgroups showed markedly different results. The Rapidly Improving group had an HR of 1.10 (95% CI: 1.01 - 1.18), suggesting a longer duration of ventilation associated with steroid use. In contrast, the Rapidly Worsening group showed an HR of 0.90 (95% CI: 0.83 - 0.97), suggesting beneficial effect of steroids, with quicker cessation of ventilation.

**Figure 5.**
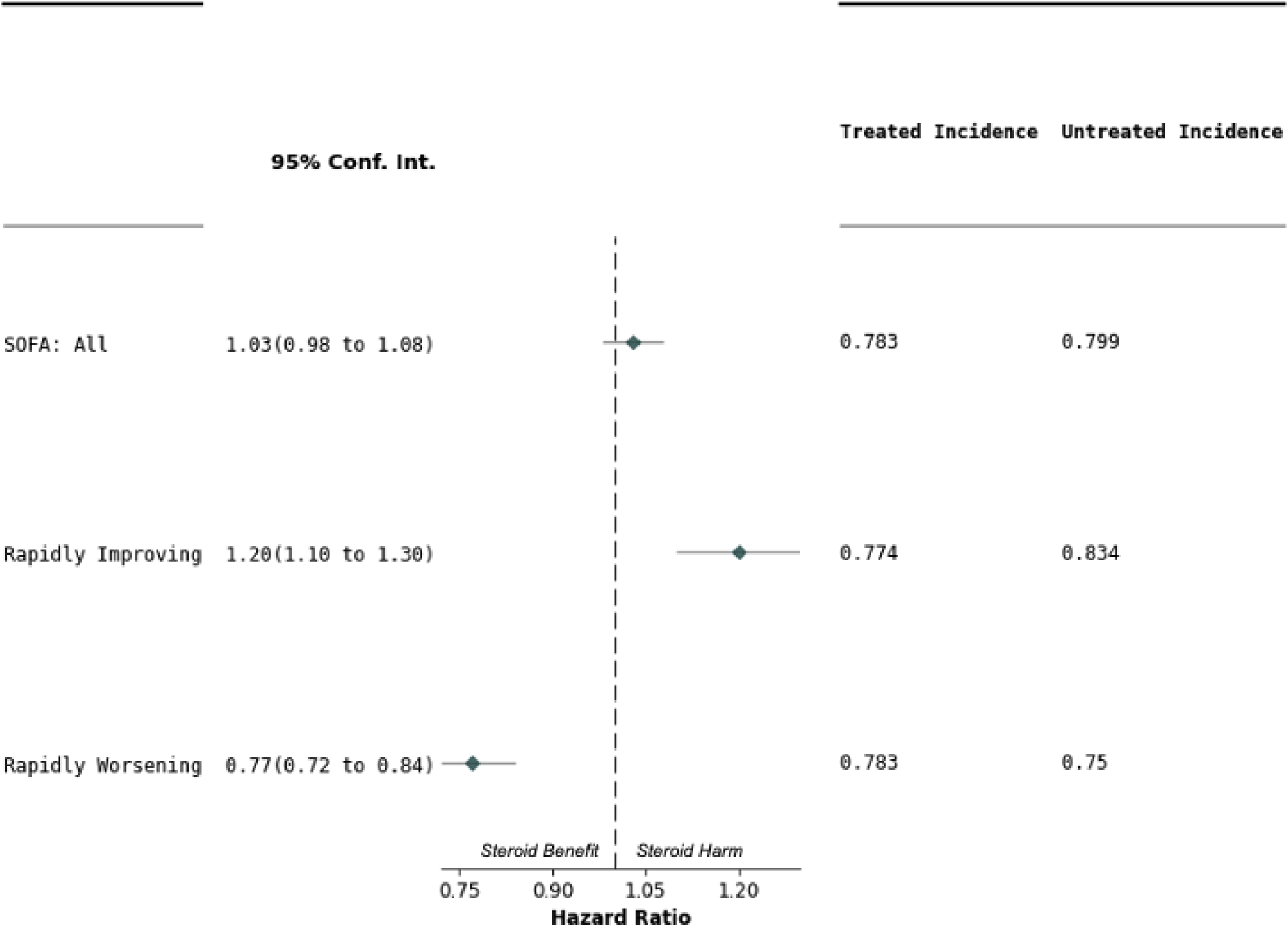
Hazard ratios and cumulative incidence for time to cessation of mechanical ventilation outcome in eICU-MIMIC cohort. Forest plot shows the hazard ratios and 95% confidence intervals for each patient stratification. Dashed vertical line represents a hazard ratio of 0. Cumulative incidences for treated and untreated populations are shown on the right.

Our sensitivity analyses (**Supplemental Figure 12 - 17**) agreed with the results of our primary approach. Across all analyses, we observed relatively increased harm for RI patients with steroid use compared to RW patients. In tandem, we observed that the incidence of 28-day mortality for RW patients was higher than that of RI patients, which is consistent with our results from the primary analysis. Our additional sensitivity analysis investigated effects of including source of infection as a covariate to our balancing method. We could only perform this analysis on a subset of the development cohort (the eICU dataset). The results show that inclusion of source of infection does not change the effects seen without source of infection significantly. We also maintain the directionality difference in terms of steroid benefit between the rapidly improving and worsening groups.

## Discussion

We examined the effects of corticosteroids in patients with infectious critical illness inclusive of ARDS, pneumonia, and septic shock, through a target trial emulation framework with real-world data from multiple hospitals. To address potential heterogeneity based on the predicted progression or resolution of organ dysfunction, we identified organ dysfunction trajectory based subphenotypes modeled on previously described methods^21^. Like our previous work, we found that the RI group was characterized by a lower in-hospital mortality rate despite a higher mean SOFA score at ICU admission (**Supplemental Table 9** and **Supplemental Figure 5**). Conversely, the RW group was characterized by metabolic acidosis (high lactate), disseminated intravascular coagulation (a higher INR and low platelets) and worse outcomes (**Supplemental Table 9** and **Supplemental Table 12-13**). Then, to evaluate the effects of corticosteroids, we predicted RI and RW group membership based on logistic regression model fit with variables available through the end of the target trial enrollment period^19^. To handle immortal time bias we set time zero for treated patients to the time of treatment and cloned all patients who died at the enrollment window^26^. We used propensity score matching to control confounding factors and reported HR for balanced emulated trials to evaluate the effectiveness of corticosteroid treatment.

In the development cohort, patients treated with steroids had different profiles from those untreated. Treated patients were younger and had a higher comorbidity burden. Both the treated and untreated groups have comparable lengths of ICU stay and baseline SOFA scores. Steroids were used more commonly in patients with pneumonia compared to patients with bacteremia. In the overall cohort our fully adjusted target trial model found that corticosteroids treatment was associated with a small increase in 28-day mortality compared to untreated controls. This mortality association was confirmed in our validation cohort. There was no average treatment effect on duration of mechanical ventilation or length of stay in the ICU.

Predicted organ dysfunction trajectory at the time of target trial enrollment was associated with a divergent treatment effect from corticosteroids. We found that there was consistent harm when steroids were used in patients with a predicted improving phenotype. The exagerated harm seen in the predicted improving phenotype may be due to several mechanisms. Improving patients with infectious critical illness may already have shifted to an adaptive immune response^40^. Treatment with corticosteroids may disrupt this process leading to inneffective clearance of the primrary infection. This is conceptually supported by a secondary analysis of the VANISH clinical trial that showed that patients with an adaptive immune transcriptomic profile were harmed by steroids compared to patients with a innate immune profile. Moreover, the risk of secondary infection, and delirium may outweigh the relative vasopressor effect in this population.

Conversely, we found a neutral treatment effect for patients with a predicted worsening phenotype in the derivation cohort and subtle harm in the validation cohort. Moreover, in patients with a predicted worsening phenotype steroid treatment was associated with shorter duration of mechanical ventilation and ICU length of stay. These results largely replicated recent large scale clinical trials of steroid use in septic shock^9^ and systematic reviews^33,34^ in a broader population. In the predicted worsening phenotype, steroids have a clear benefit in time to organ dysfunction resolution, reported in multiple clinical trials and this target trial study supports this effect^35,36^.

The finding that steroids benefit patients who are predicted to have worsening organ dysfunction is consistent with the proposed biologic mechanism for the action of steroids in critically ill patients^37^. Specifically, the rapidly worsening phenotype may benefit from the vasopressor effect of systemic corticosteroid to improve tissue perfusion and ameliorate visceral organ dysfunction. The steroid effect on immune cell death through activation of the apoptotic pathways may help limit the disregulated innate inflammatory response and potentially disrupt the beneficial adaptive immune response. These hypotheses should be tempered, however, by the existing evidence that the adrenalcortical candidate gene expression did not predict steroid response in a randomized clinical trials^15,38^. Moreover, traditional markers of suppression of the hypothalamic pituitary axis such as serum cortisol, do not reliably predicte the response to corticosteroids^39^.

Our findings can provide several implications for clinicians and other researchers. Firstly, we identified differential average treatment effects based on different predicted organ function trajectory. The findings reinforce the concept that disease states are dynamic, and researchers may have to consider the response to standard treatment when considering drug effectiveness in heterogeneous diseases. Secondly, our ML based stratification method is based on predictive modeling, which can facilitate early interventions before disease trajectory is established and would have the potential to lead to a significant impact on trial enrollment. Thirdly, our study regarding the evaluation of the effectiveness of corticosteroids in patients with sepsis was conducted through a trial-emulated framework with real-world data, which could be important complementary evidence to the findings of RCTs^9,41^.

This study has several limitations. We used a modification of the Sepsis-3 framework to identify patients for this study examining the trajectory of organ dysfunction. The individual components of the SOFA score may not fully capture all infectious related critical illness. Moreover, the study’s observational design implies that despite rigorous propensity score matching, residual confounding cannot be completely ruled out. Although we made significant efforts to balance confounders across treatment groups, some unmeasured or unknown confounders might still influence the observed results. To measure the influence of these unmeasured confounders, we calculated E-values on our effects using the formula developed by Tyler J. VanderWeele^42^. These E-values suggest the effects of unmeasured confounders is minimal (**Supplemental Table 14, Supplemental Text 2**). Second, we used real-world data from ICU databases, which may be subject to inherent biases such as missing data, measurement errors, and variability in data collection practices across different institutions. However, we believe it is valuable to use this real-world data to determine the effect of treatments in specific clinical cohorts. Third, our machine learning approach, while revealing meaningful patient subphenotypes, relies on certain assumptions. For example, we assume that the SOFA score trajectories effectively capture the underlying patient states, which may not fully encompass the complexity of organ failure progression. That said, different outcomes and characteristics were observed between our predicted groups that fit the model of differential progression. Lastly, the performance of our logistic regression models for predicting subphenotypes based on baseline covariates could have been better, leading to more heterogeneous differences between the RI and RW groups. Discrimination between RI and RW groups could be enhanced through testing steroid responsive transcriptomics or protein pathway enrichment by group, enabling it to generalize to ICUs with different populations.

## Conclusion

Our study provides a nuanced view of the role of corticosteroids in sepsis management, in several overlapping syndromic disease states. We identified differential average treatment effects when stratified by trajectory based subphenotypes. This finding highlights the potential for personalized treatment strategies in sepsis, informed by subphenotype rather than syndrome. Future research should focus on validating these machine learning subphenotypes in independent cohorts, in randomized clinical trials as well as exploring their biological basis.

## Data Availability

The de-identified data utilized in this study for the development cohort (MIMIC-IV) and the validation cohort (eICU) can be accessed upon the approval of a formal proposal and the execution of a Data Access Agreement via Physio Net (https://physionet.org/). Access for the CEDAR validation cohort is not encompassed by our existing data transfer agreements.

## Code Availability

The source code pertinent to this research is publicly accessible. The primary repository is hosted on https://github.com/surajraj99/Corticosteroids-in-Patients-with-Sepsis. Source code for determining subtypes can be found in https://github.com/xuzhenxing2019/sepsis_subphenotype.

## Acknowledgements

The authors would like to acknowledge the support from National Science Foundation awards (nos. 1750326 and 2212175), National Institute of Health awards (nos. RF1AG072449, R01AG080624, R01AG076448, RF1AG084178, R01AG076234, R01AG080991 and R01MH 124740), National Institute of Health award NHLBI K23 HL151876-01A1, and National Institute of General Medical Sciences award number K23 GM151730-01 for this study.

## Author Contributions

S.R., E.S., F.W., and Z.X. conceived the study. S.R., E.S., Z.X., W.P. and C.Z. conceived the method and designed the algorithmic techniques. S.R. wrote the codes and performed the computational analysis with input from Z.X, W.P., C.Z. All the authors read the paper and suggested edits. F.W. supervised the project.

