## Supplemental Data for "Corticosteroids for infectious critical illness: A multicenter target trial emulation stratified by predicted organ dysfunction trajectory"

**Supplemental Information**

**S1. Supplemental Tables**

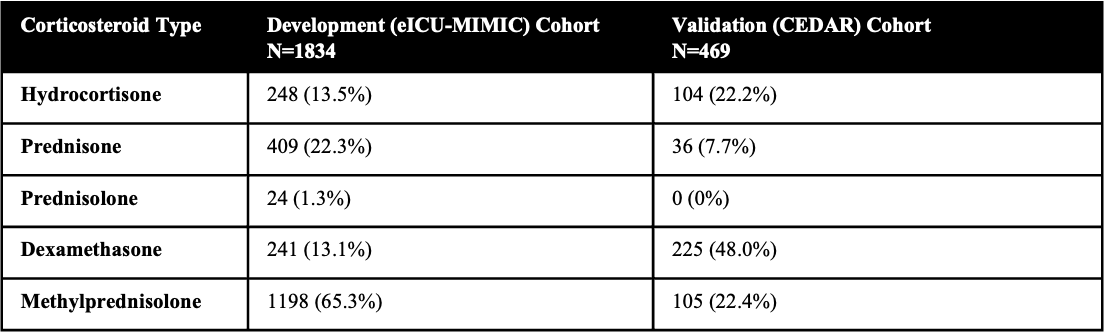

**Supplemental Table 1. Breakdown of corticosteroid subtype used by treated patients.** Absolute counts as well as the percentage of treated patients who were treated with the specific corticosteroid subtype are shown for both development and validation cohort.

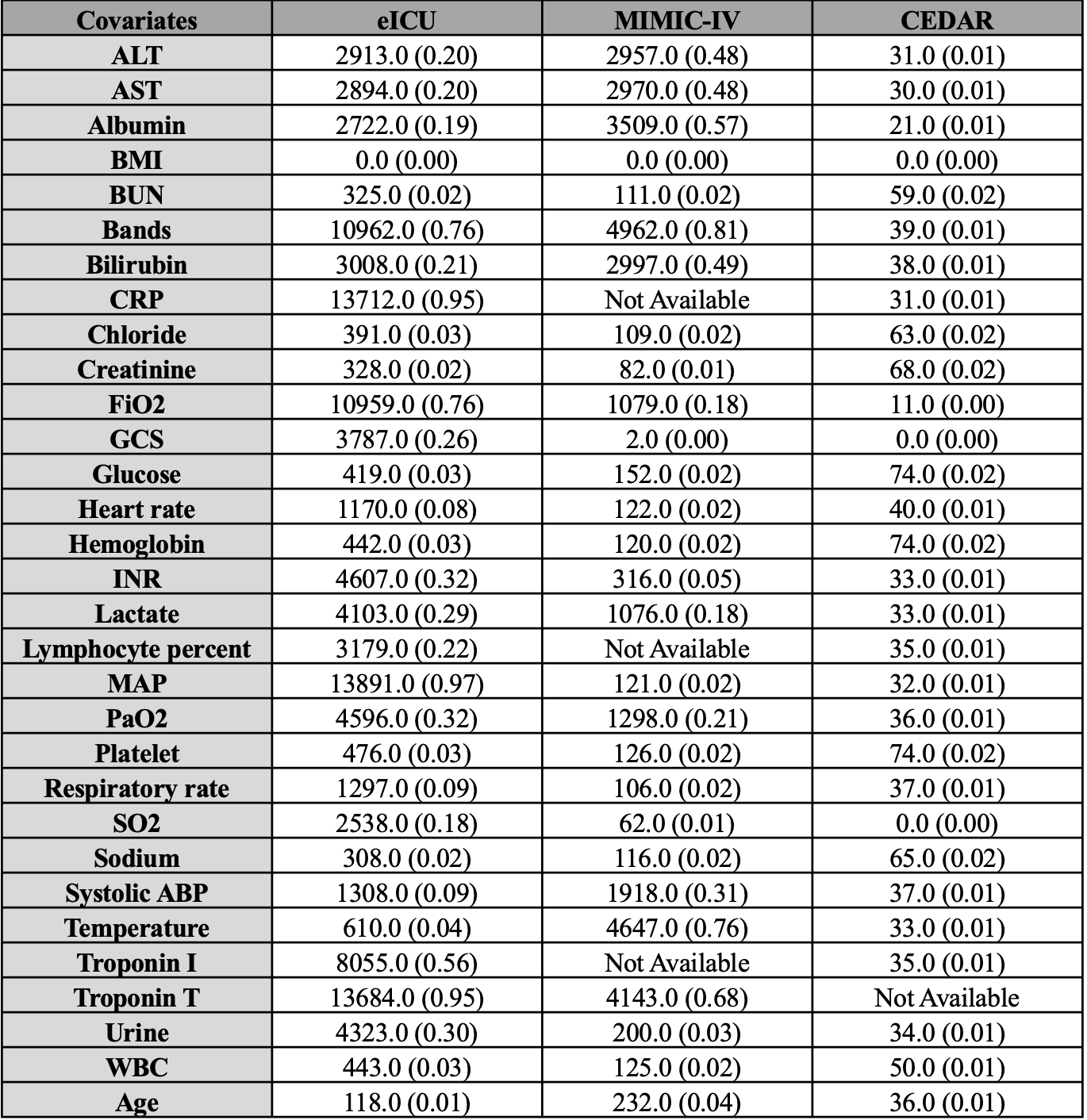

**Supplemental Table 2**. **Missingness of covariates within each data cohort**. Parentheses denote the percentage of covariates missing within the cohort.

| **Protocol component** | **Target trial specification** | **Target trial emulation** |
| --- | --- | --- |
| **Eligibility criteria** | Age >=18 at enrollment window  Patient with sepsis at enrollment window  No sepsis history before enrollment window  No corticosteroid prescription more than 10 hrs before enrollment window  Sepsis is defined based on Sepsis-3 criteria  The enrollment window is defined as 24 hours after the ICU admission [0h, 24h] | Age >=18 at enrollment window  We require the first ICU stay if he/she has multiple ICU stays in the database  No sepsis history before enrollment window  We define sepsis based on Sepsis-3 with the record at his/her enrollment window  The enrollment window is defined as the first day of the patient’s ICU admission |
| **Treatment strategies** | Strategy 0: No initiation of any corticosteroids drug at enrollment window    Strategy 1: Initiation of hydrocortisone at a dose of 160 mg at enrollment window | We compute cumulative milligram dosing of hydrocortisone at enrollment window, and if a patient received more than 160 mg per day hydrocortisone equivalent, they are denoted as having hydrocortisone exposure.    We consider corticosteroids including Prednisolone, Prednisone, Hydrocortisone, Dexamethasone, and Methylprednisolone.    **Conversions:**  1 mg Prednisolone = 4 mg Hydrocortisone  1 mg Prednisone = 4 mg Hydrocortisone  1 mg Dexamethasone =  25 mg Hydrocortisone  1 mg Methylprednisolone = 5 mg Hydrocortisone |
| **Treatment assignment** | Patients are randomly assigned to either treatment strategy on the first day after ICU admission and are aware of the strategy they are assigned to. | We classified individuals according to the  strategy that their data were compatible with at the enrollment window and attempted to emulate  randomization by adjusting for baseline  confounders. |
| **Outcomes** | 28-day mortality from time of randomization | Same as for the target trial. |
| **Follow-up** | We follow each patient from his/her baseline until the day of his/her death, loss to follow-up, or discharge, whichever occurs first. | Same as for the target trial. |
| **Causal contrasts** | Intention-to-treat effect | Observational analog of intention-to-treat effect |
| **Statistical analysis** | We use Kaplan-Meier survival plot to report the rate of death in a time-to-event analysis.  We use Cox proportional hazards to report the differences in survival. | Same as for the target trial.    To model the cumulative incidence function, we assign individuals who discharge the "best" outcome (right-censoring them at the longest recorded LOS or TCS)    Patients whose transfer location are unknown are censored at time transfer |

**Supplemental Table 3**. **Target trial emulation for 28-day mortality outcome**.

| **Protocol component** | **Target trial specification** | **Target trial emulation** |
| --- | --- | --- |
| **Eligibility criteria** | Age >=18 at enrollment window  Patient with sepsis at enrollment window  No sepsis history before enrollment window  No corticosteroid prescription more than 10 hrs before enrollment window  Sepsis is defined based on Sepsis-3 criteria  The enrollment window is defined as 24 hours after the ICU admission [0h, 24h] | Age >=18 at enrollment window  We require the first ICU stay if he/she has multiple ICU stays in the database  No sepsis history before enrollment window  We define sepsis based on Sepsis-3 with the record at his/her enrollment window  The enrollment window is defined as the first day of the patient’s ICU admission |
| **Treatment strategies** | Strategy 0: No initiation of any corticosteroids drug at enrollment window    Strategy 1: Initiation of hydrocortisone at a dose of 160 mg at enrollment window | We compute cumulative milligram dosing of hydrocortisone at enrollment window, and if a patient received more than 160 mg per day hydrocortisone equivalent, they are denoted as having hydrocortisone exposure.    We consider corticosteroids including Prednisolone, Prednisone, Hydrocortisone, Dexamethasone, and Methylprednisolone. |
| **Treatment assignment** | Patients are randomly assigned to either treatment strategy on the first day after ICU admission and are aware of the strategy they are assigned to. | We classified individuals according to the  strategy that their data were compatible  with at the enrollment window and attempted to emulate  randomization by adjusting for baseline  confounders. |
| **Outcomes** | Time to ICU Discharge from time of randomization | Same as for the target trial. |
| **Follow-up** | We follow each patient from his/her baseline until the day of his/her death, loss to follow-up, or discharge, whichever occurs first. | Same as for the target trial. |
| **Causal contrasts** | Intention-to-treat effect | Observational analog of intention-to-treat effect |
| **Statistical analysis** | We use Kaplan-Meier survival plot to report the rate of death in a time-to-event analysis.  We use Cox proportional hazards to report the differences in survival. | Same as for the target trial.    To model the cumulative incidence function, we assign individuals who die the "worst" outcome (right-censoring them at the longest recorded LOS or TCS)    Patients whose transfer location are unknown are censored at time of transfer |

**Supplemental Table 4**. **Target trial emulation for time to ICU discharge outcome**.

| **Protocol component** | **Target trial specification** | **Target trial emulation** |
| --- | --- | --- |
| **Eligibility criteria** | Age >=18 at enrollment window  Patient with sepsis at enrollment window  No sepsis history before enrollment window  No corticosteroid prescription more than 10 hrs before enrollment window  Sepsis is defined based on Sepsis-3 criteria  The enrollment window is defined as 24 hours after the ICU admission [0h, 24h]  On mechanical ventilation at enrollment window | Age >=18 at enrollment window  We require the first ICU stay if he/she has multiple ICU stays in the database  No sepsis history before enrollment window  We define sepsis based on Sepsis-3 with the record at his/her enrollment window  The enrollment window is defined as the first day of the patient’s ICU admission  On mechanical ventilation at enrollment window |
| **Treatment strategies** | Strategy 0: No initiation of any corticosteroids drug at enrollment window    Strategy 1: Initiation of hydrocortisone at a dose of 160 mg at enrollment window | We compute cumulative milligram dosing of hydrocortisone at enrollment window, and if a patient received more than 160 mg per day hydrocortisone equivalent, they are denoted as having hydrocortisone exposure.    We consider corticosteroids including Prednisolone, Prednisone, Hydrocortisone, Dexamethasone, and Methylprednisolone. |
| **Treatment assignment** | Patients are randomly assigned to either treatment strategy on the first day after ICU admission and are aware of the strategy they are assigned to. | We classified individuals according to the  strategy that their data were compatible  with at the enrollment window and attempted to emulate  randomization by adjusting for baseline  confounders. |
| **Outcomes** | Time to ventilation cessation from time of randomization, ventilation cessation is defined as 24 hours of no ventilation support | Same as for the target trial. |
| **Follow-up** | We follow each patient from his/her baseline until the day of his/her death, loss to follow-up, or discharge, whichever occurs first. | Same as for the target trial. |
| **Causal contrasts** | Intention-to-treat effect | Observational analog of intention-to-treat effect |
| **Statistical analysis** | We use Kaplan-Meier survival plot to report the rate of death in a time-to-event analysis.  We use Cox proportional hazards to report the differences in survival. | Same as for the target trial.    To model the cumulative incidence function, we assign individuals who die the "worst" outcome (right-censoring them at the longest recorded LOS or TCS)    Patients who are discharged are marked as ceasing ventilation at time of discharge if ventilation cessation was not marked earlier    Patients whose transfer location are unknown are censored at time of transfer |

**Supplemental Table 5**. T**arget trial emulation for cessation of mechanical ventilation outcome**.

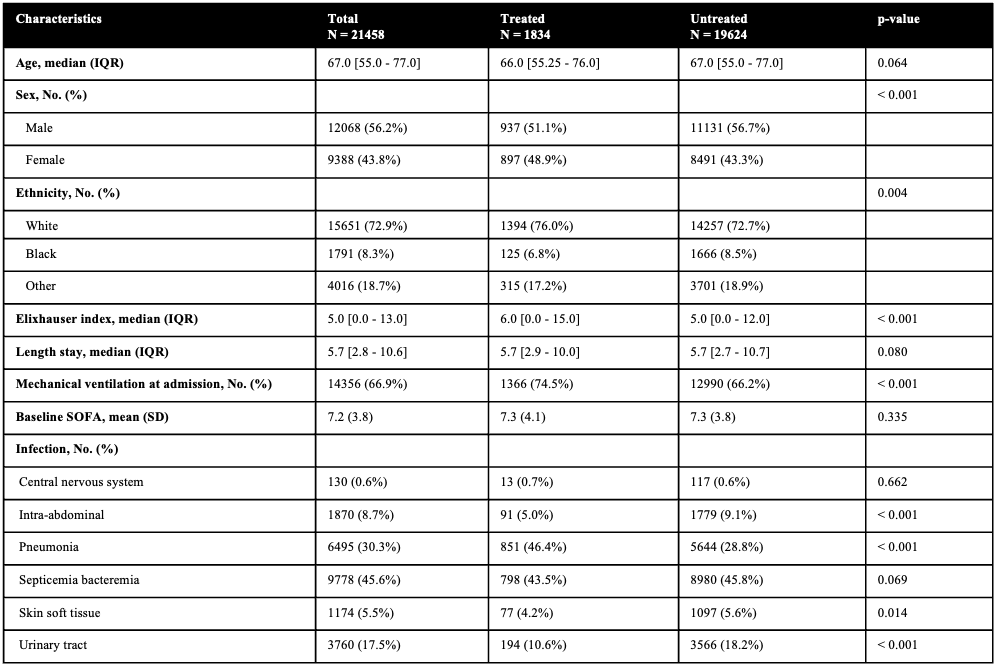

**Supplemental Table 6. Patient characteristics comparisons between treated and untreated in the eICU-MIMIC cohort.** p-value(s) are determined by either Chi-square exact or student's t-test where applicable.

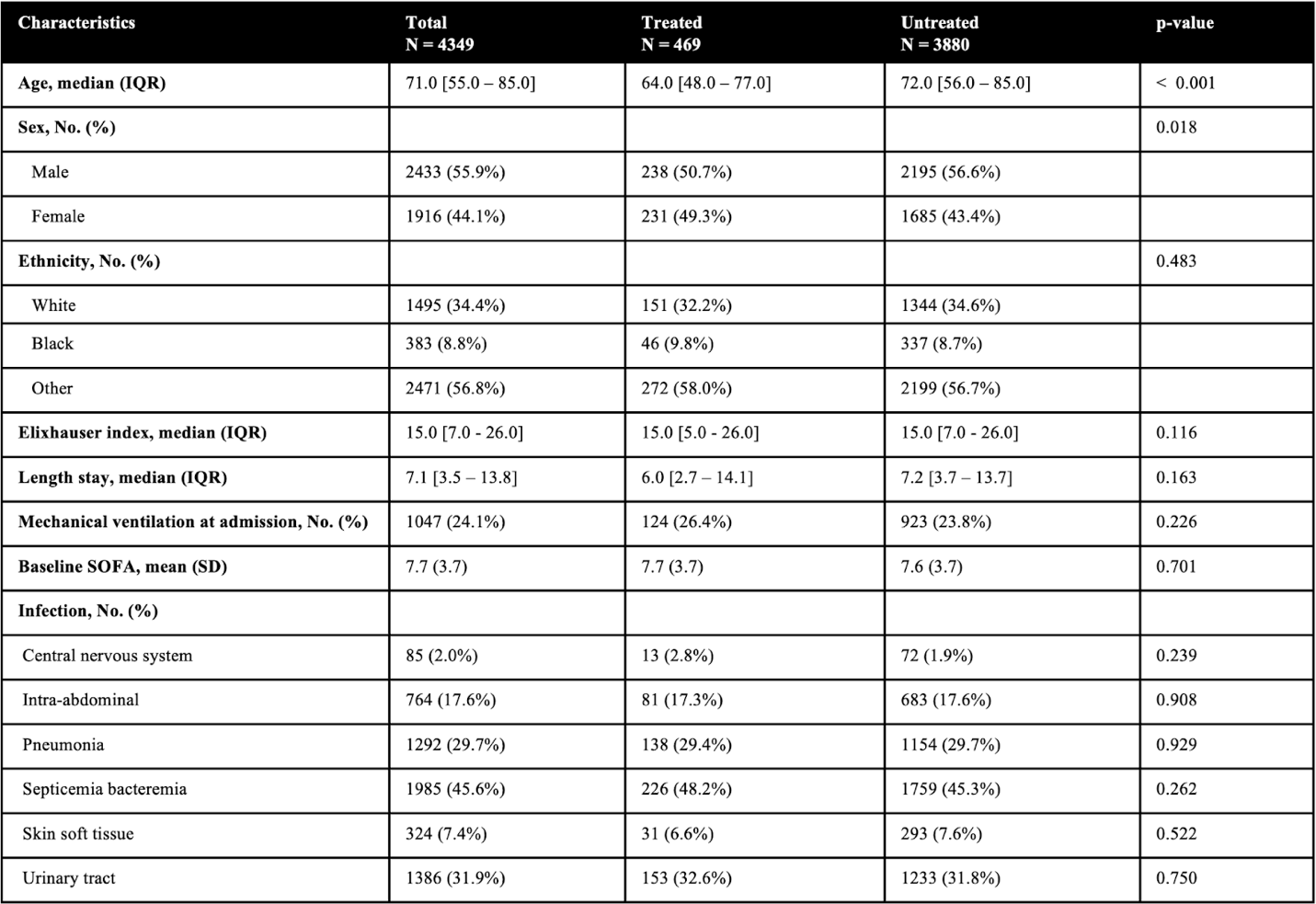

**Supplemental Table 7. Patient characteristics comparisons between treated and untreated in the CEDAR cohort.** p-value(s) are determined by either Chi-square exact or student's t-test where applicable.

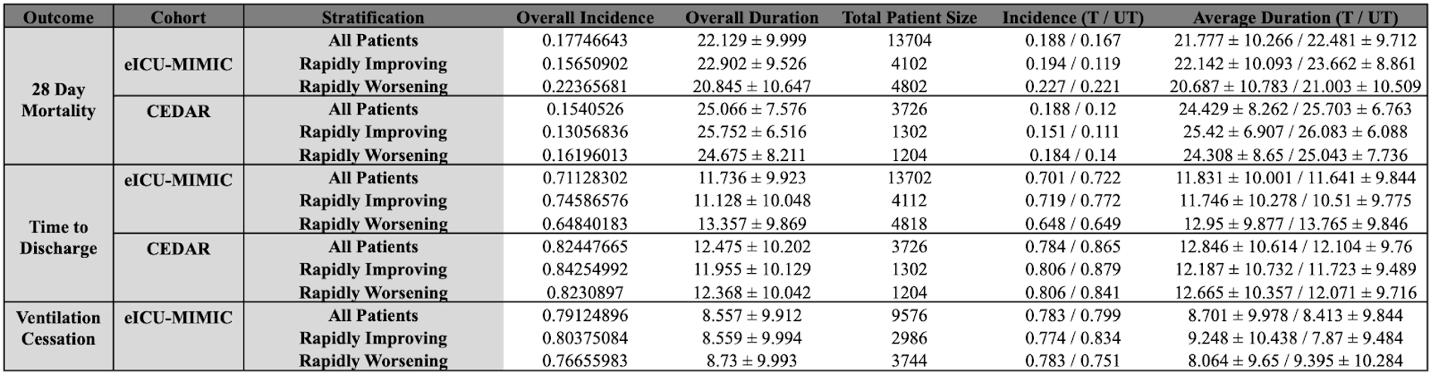

**Supplemental Table 8. Incidence of events and duration of patient stays across cohorts, outcomes, and stratification.** Shows the statistics of treated and untreated patients after performing balancing via propensity score matching.

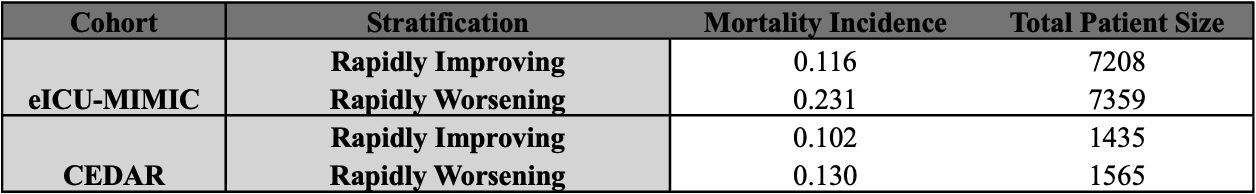

**Supplemental Table 9. Mortality incidence of machine learning subgroups in both cohorts.** Shows incidence before balancing confounders.

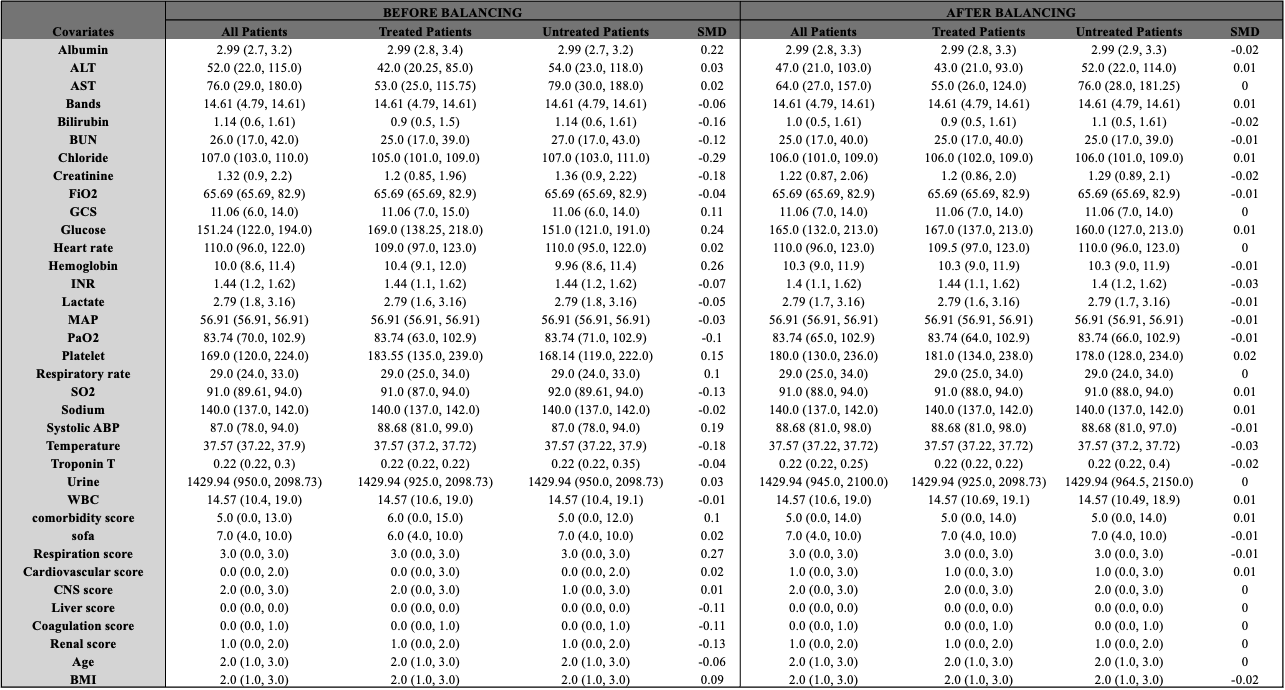

**Supplemental Table 10**. Measurements for eICU-MIMIC cohort before and after balancing for SOFA All 28-day mortality and time-to-discharge outcomes. Median and IQR values are shown for all covariates.

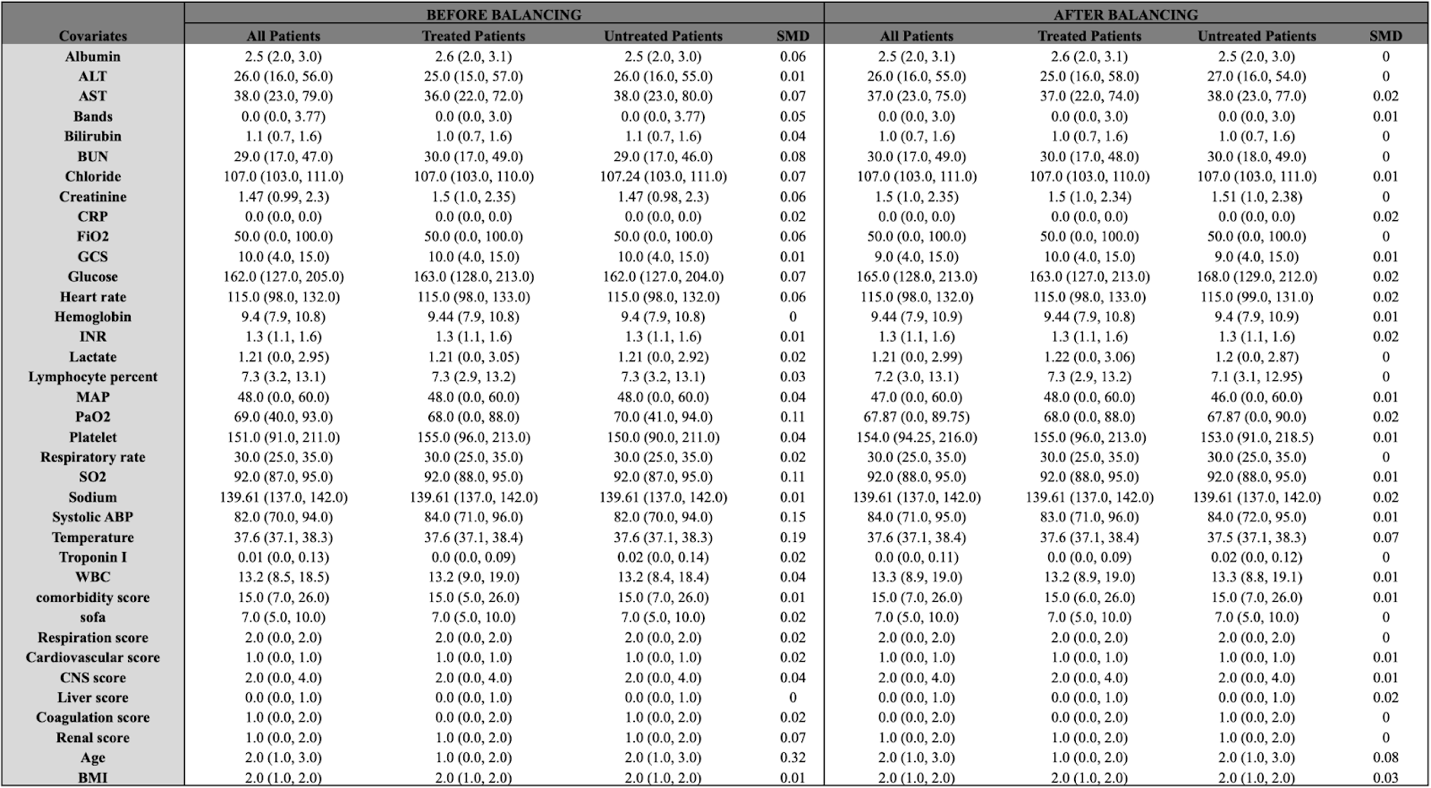

**Supplemental Table 11**. Measurements for CEDAR cohort before and after balancing for SOFA All 28-day mortality and time-to-discharge outcomes. Median and IQR values are shown for all covariates.

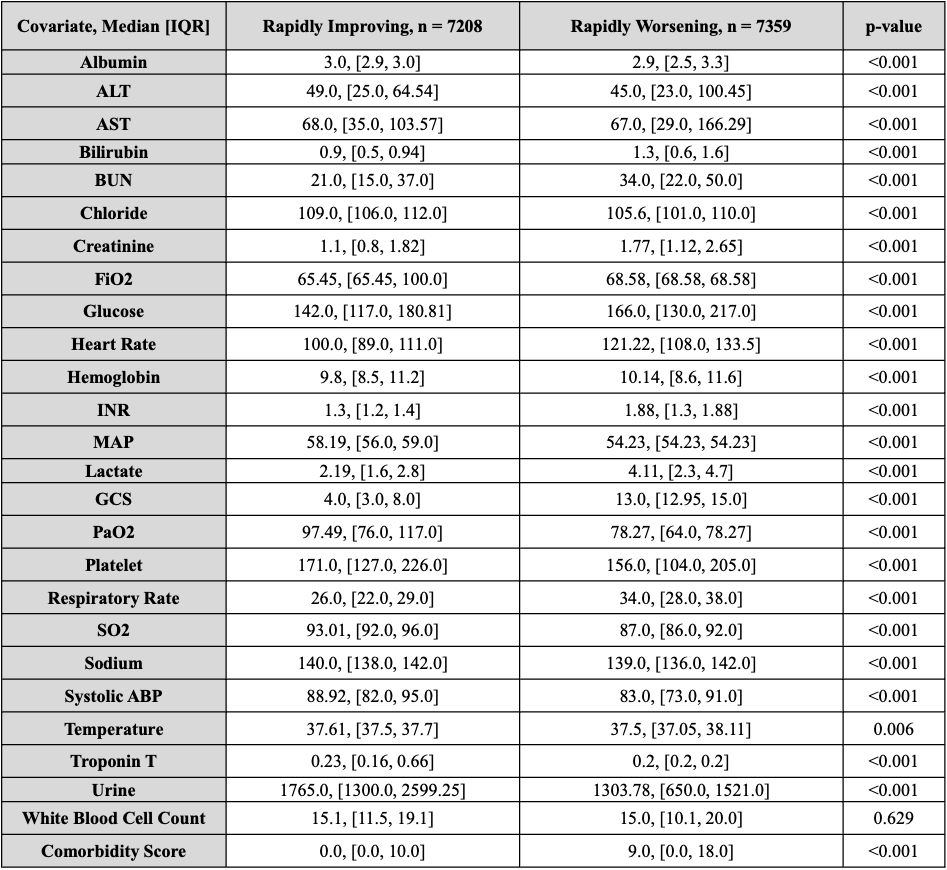

**Supplemental Table 12. Patient characteristics comparisons between Rapidly Improving and Rapidly Worsening in the eICU-MIMIC cohort.** Median and IQR is shown for select covariates. P-value(s) are determined by student's t-test.

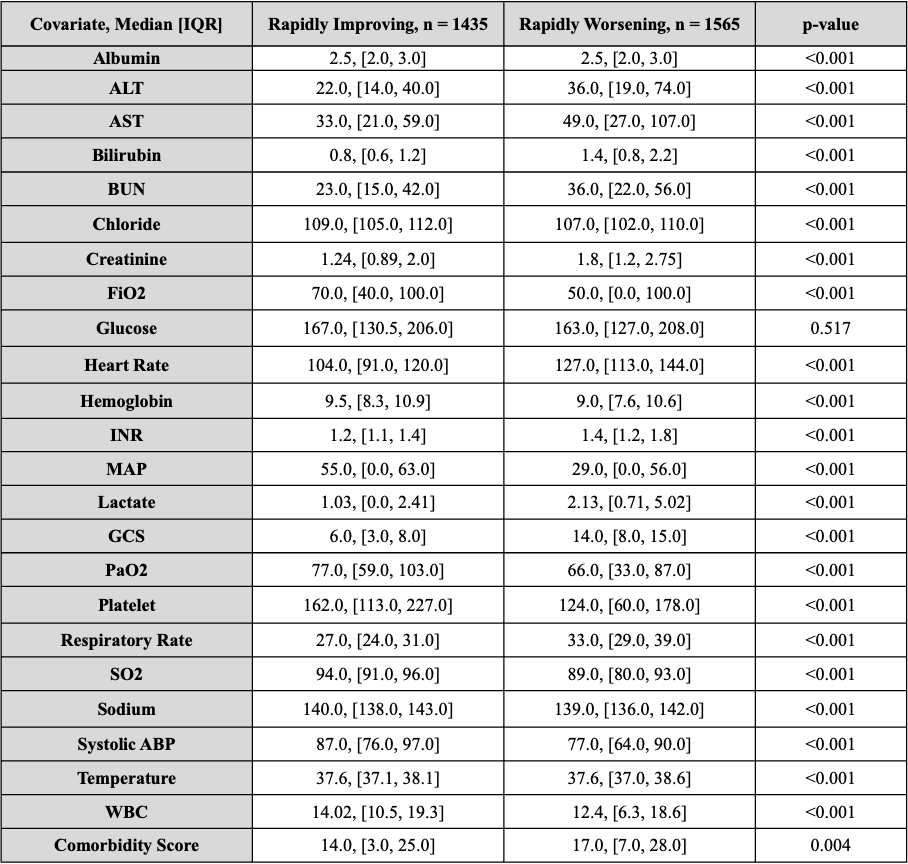

**Supplemental Table 13. Patient characteristics comparisons between Rapidly Improving and Rapidly Worsening in the CEDAR cohort.** Median and IQR is shown for select covariates. P-value(s) are determined by student's t-test.

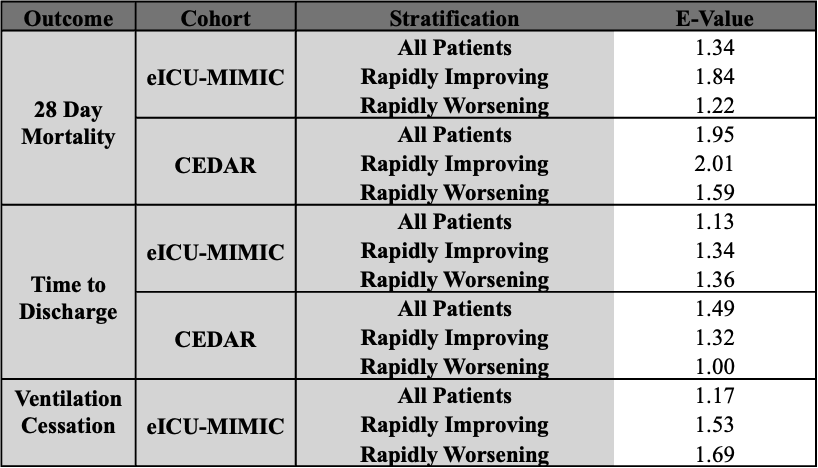

**Supplemental Table 14.** **E-values for primary and secondary analyses in development and validation cohort.** E-values were computed using the  E-value formula developed by Tyler J. VanderWeele. E-values are shown for each stratification.

**S2. Supplemental Figures**

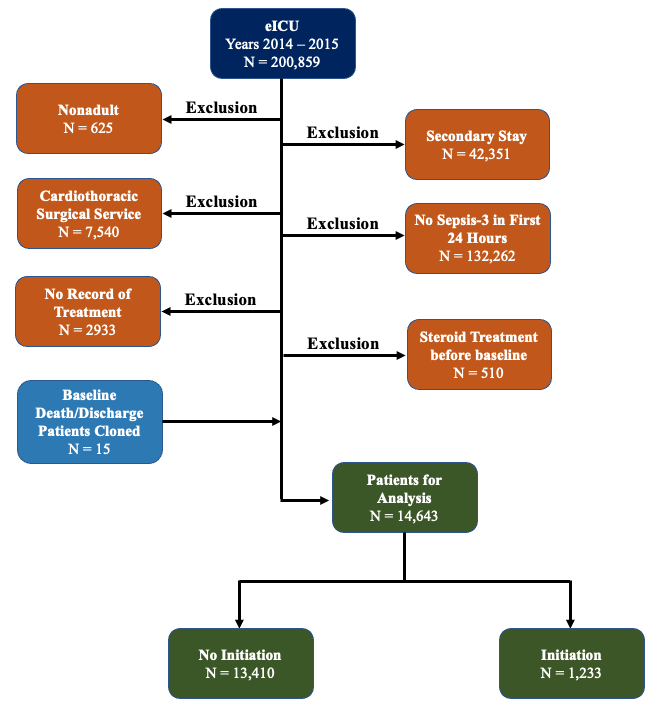

**Supplemental Figure 1**: **eICU Cohort Selection flowchart.**

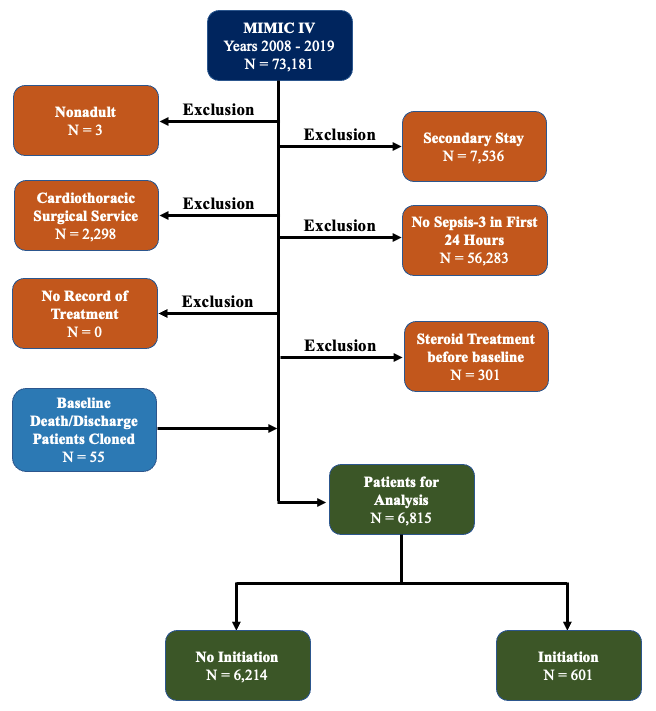

**Supplemental Figure 2**: **MIMIC-IV Cohort Selection flowchart.**

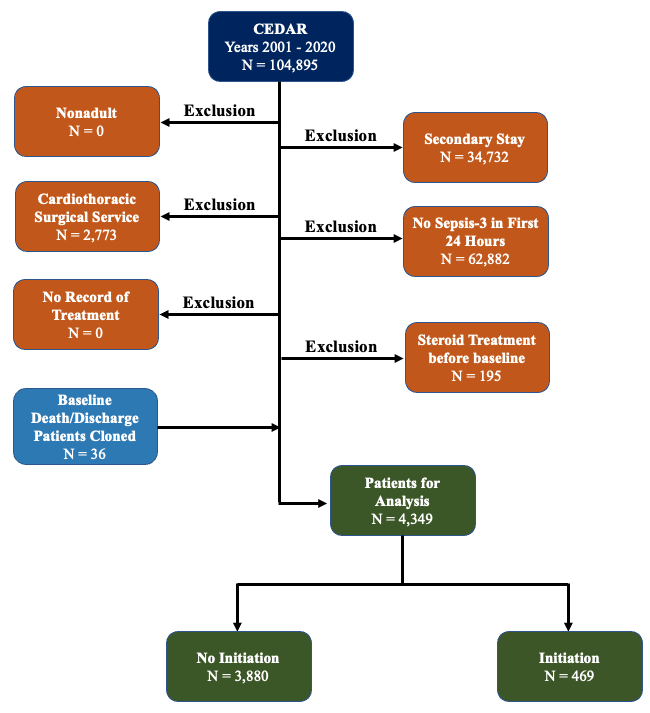

**Supplemental Figure 3**: **CEDAR Cohort Selection flowchart.**

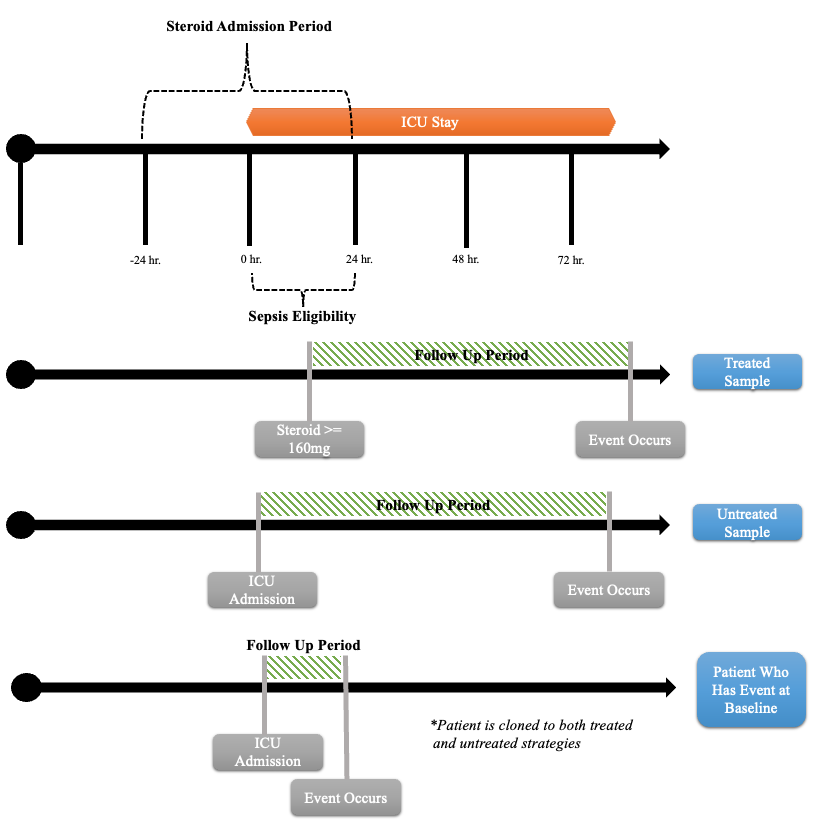

**Supplemental Figure 4. Eligibility criteria and follow up timeline.** Three sample patients are shown, a treated patient, an untreated patient, and a patient who dies before enrollment window for steroid admission. Follow-up period is shown in green. For patients who die before end of enrollment window (Day 1), they are cloned to both treated and untreated strategies.

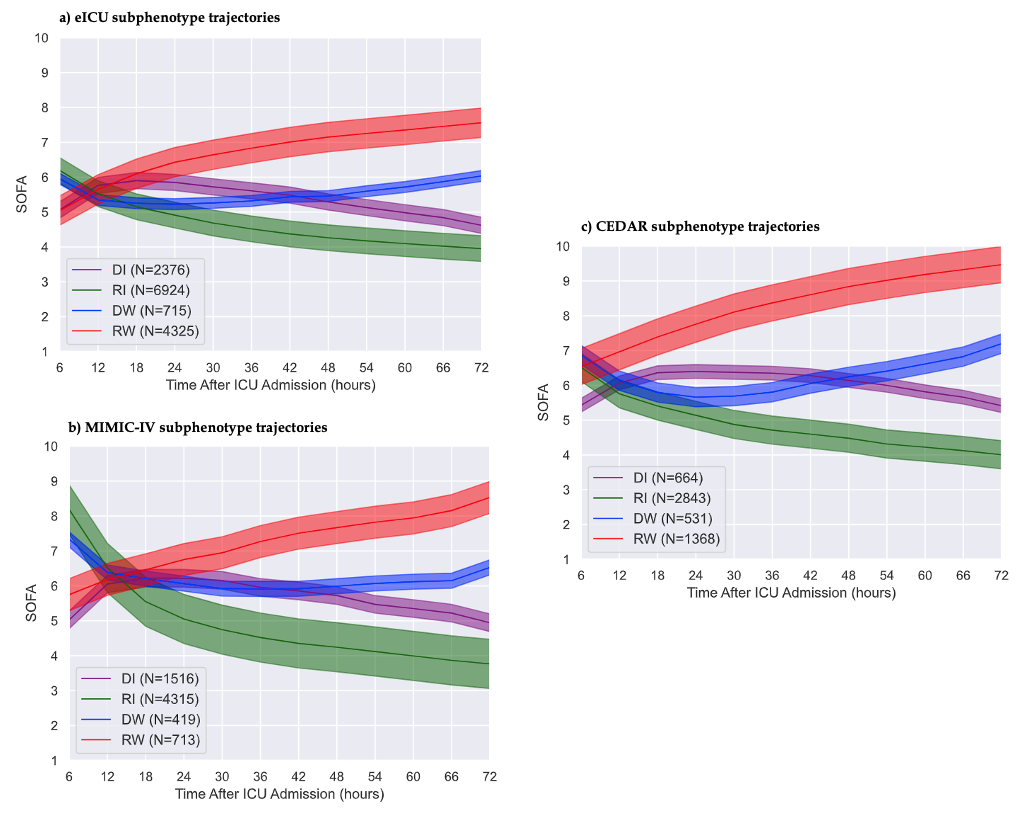

**Supplemental Figure 5**. **SOFA trajectories for machine learning subtypes across study cohorts.** Only RI (green) and RW (red) were considered in this study due to sample size constraints. (a) shows SOFA trajectories for eICU cohort, (b) shows trajectories for MIMIC-IV cohort, (c) shows trajectories for CEDAR cohort.

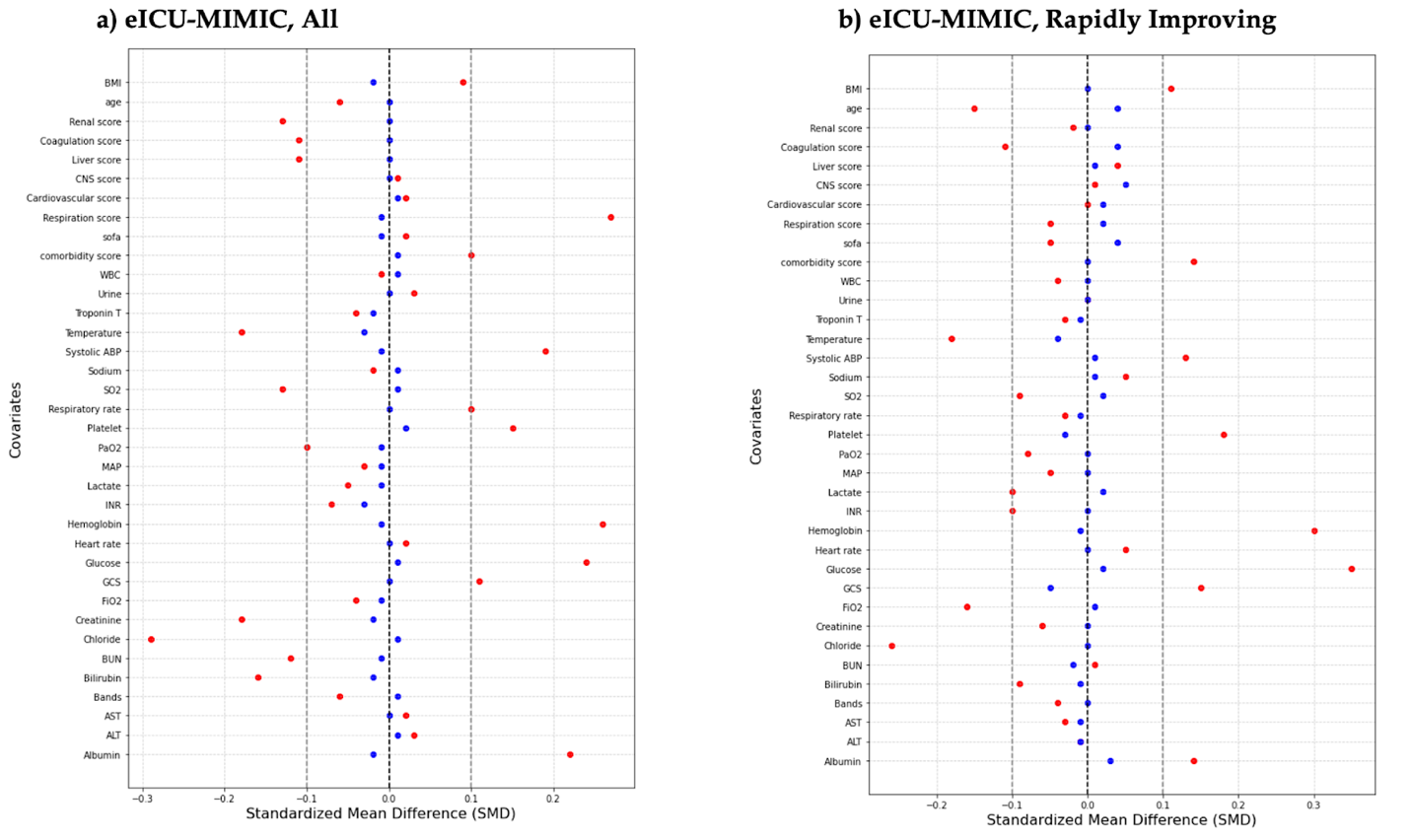

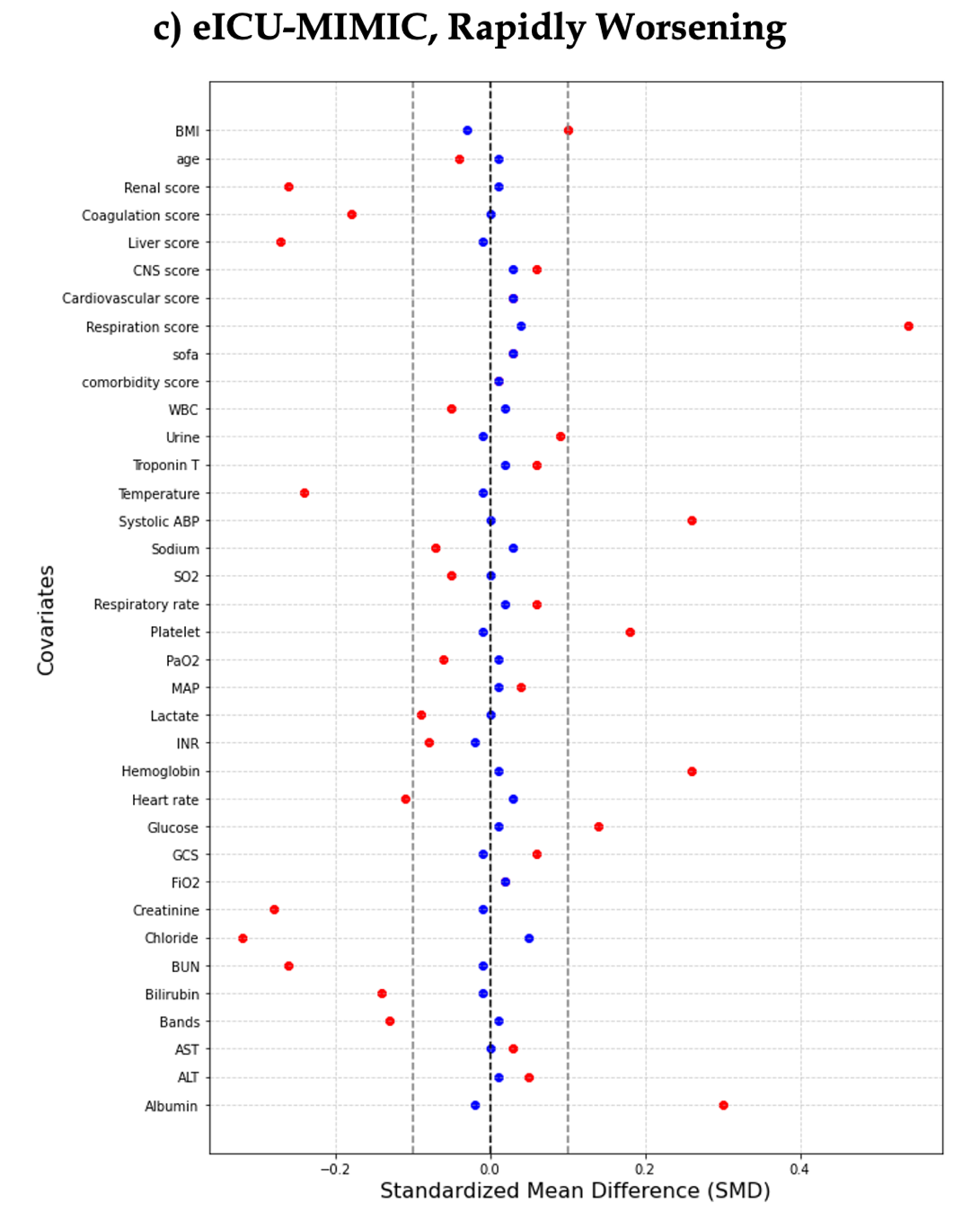

**Supplemental Figure 6**: SMD of covariates in eICU-MIMIC cohort before and after propensity score matching. SMDs are shown for 28-day mortality and time-to-discharge outcomes. Red dots indicate SMD before matching.

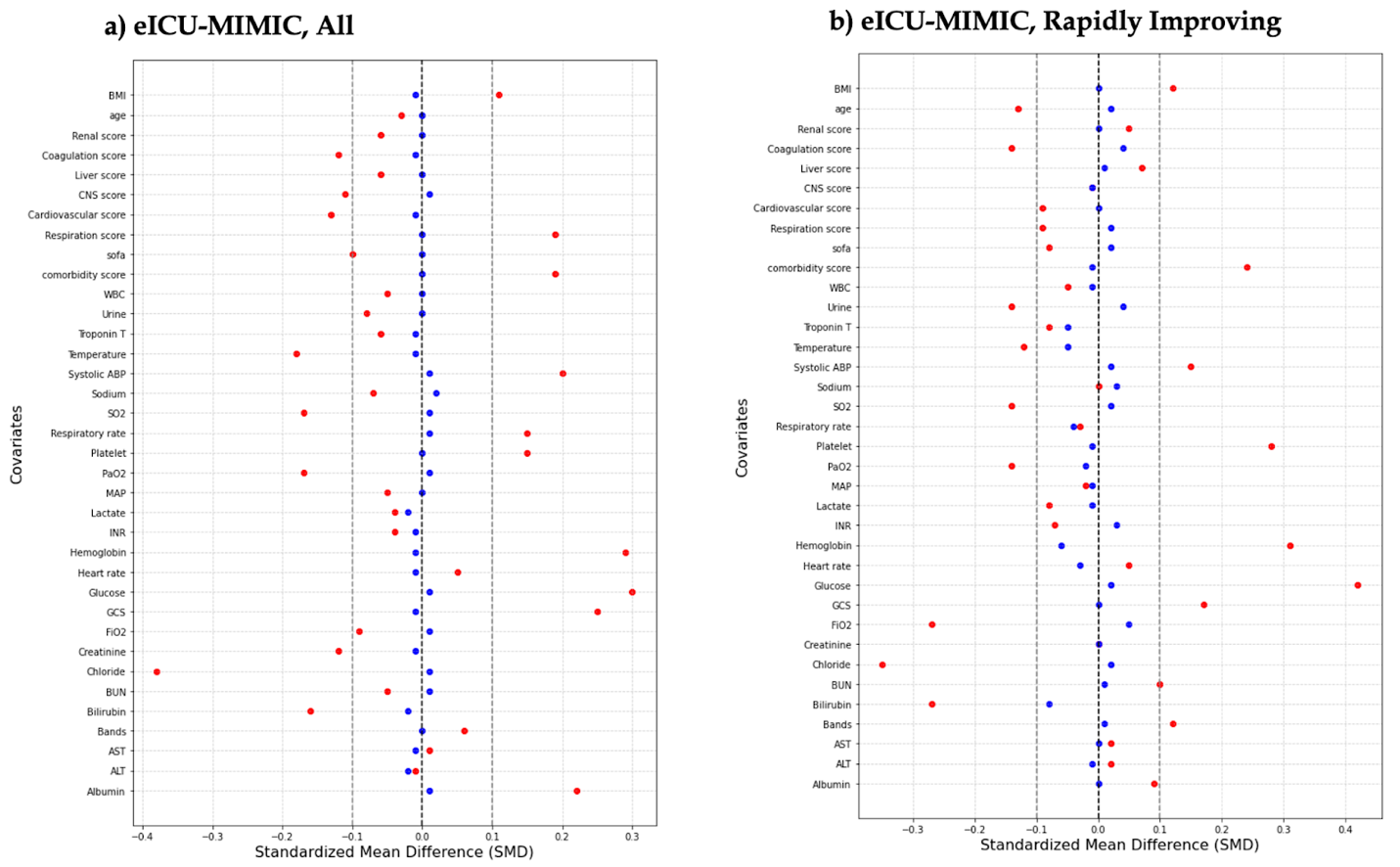

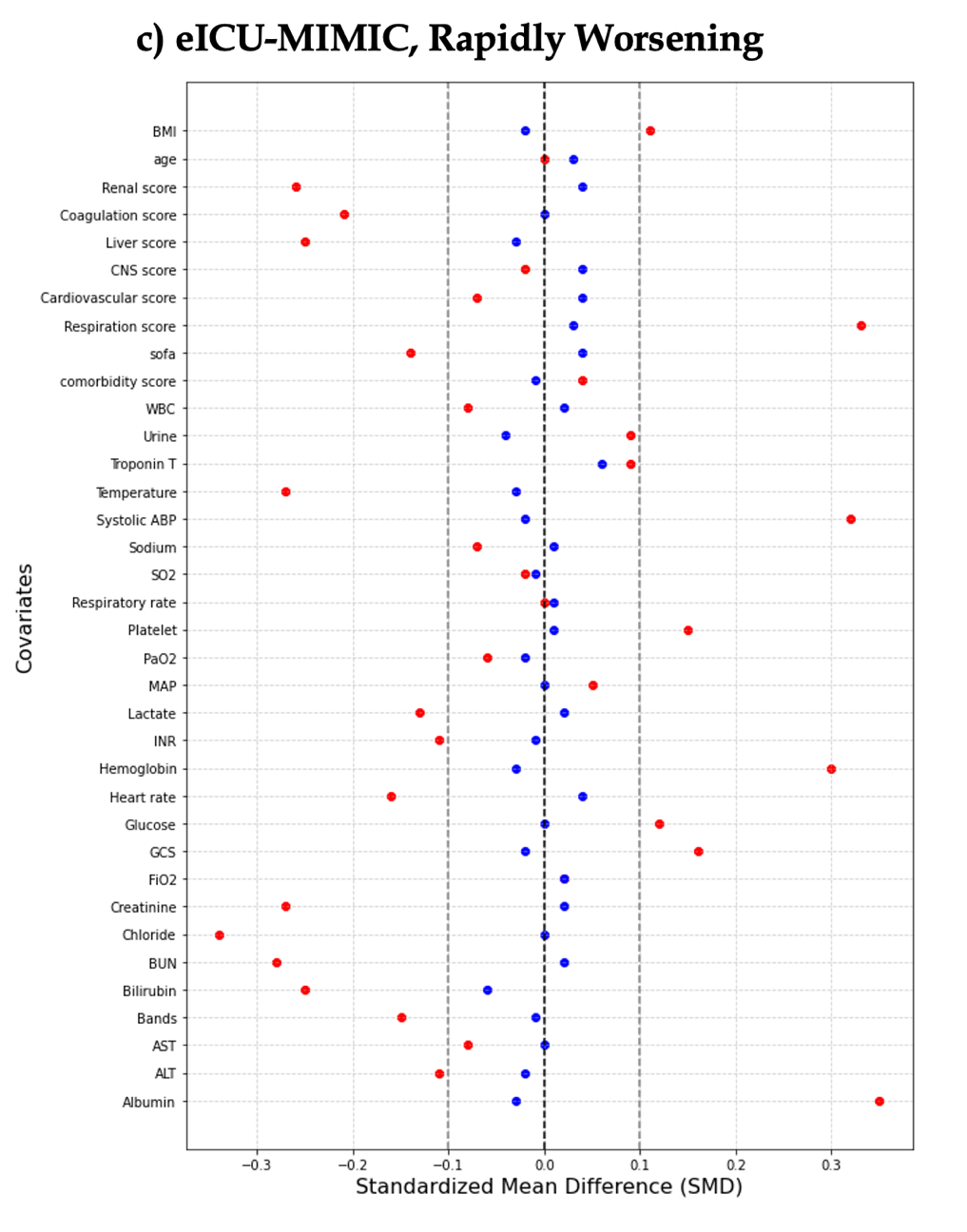

**Supplemental Figure 7**: SMD of covariates in eICU-MIMIC cohort before and after propensity score matching. SMDs are shown for ventilation cessation. Red dots indicate SMD before matching.

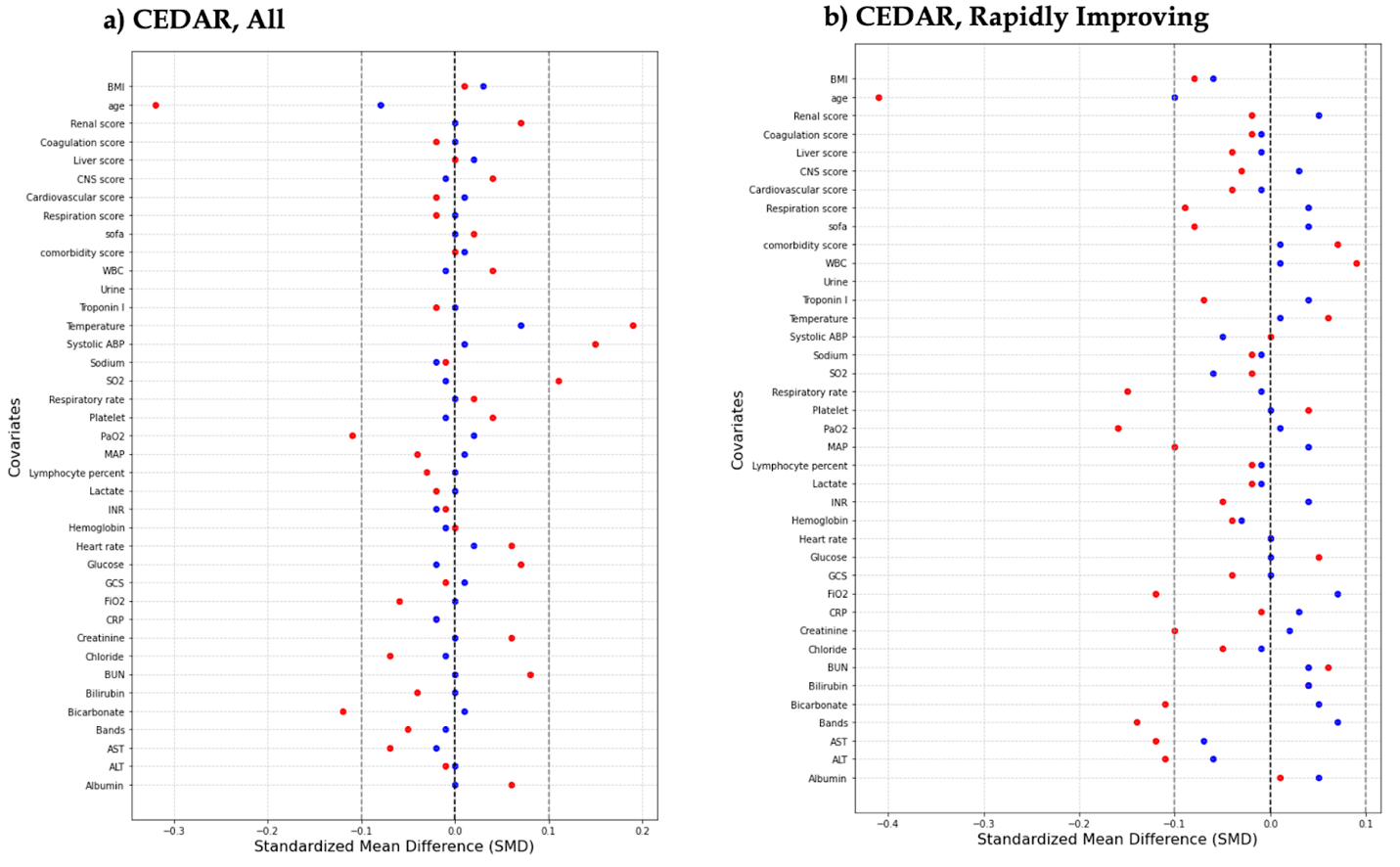

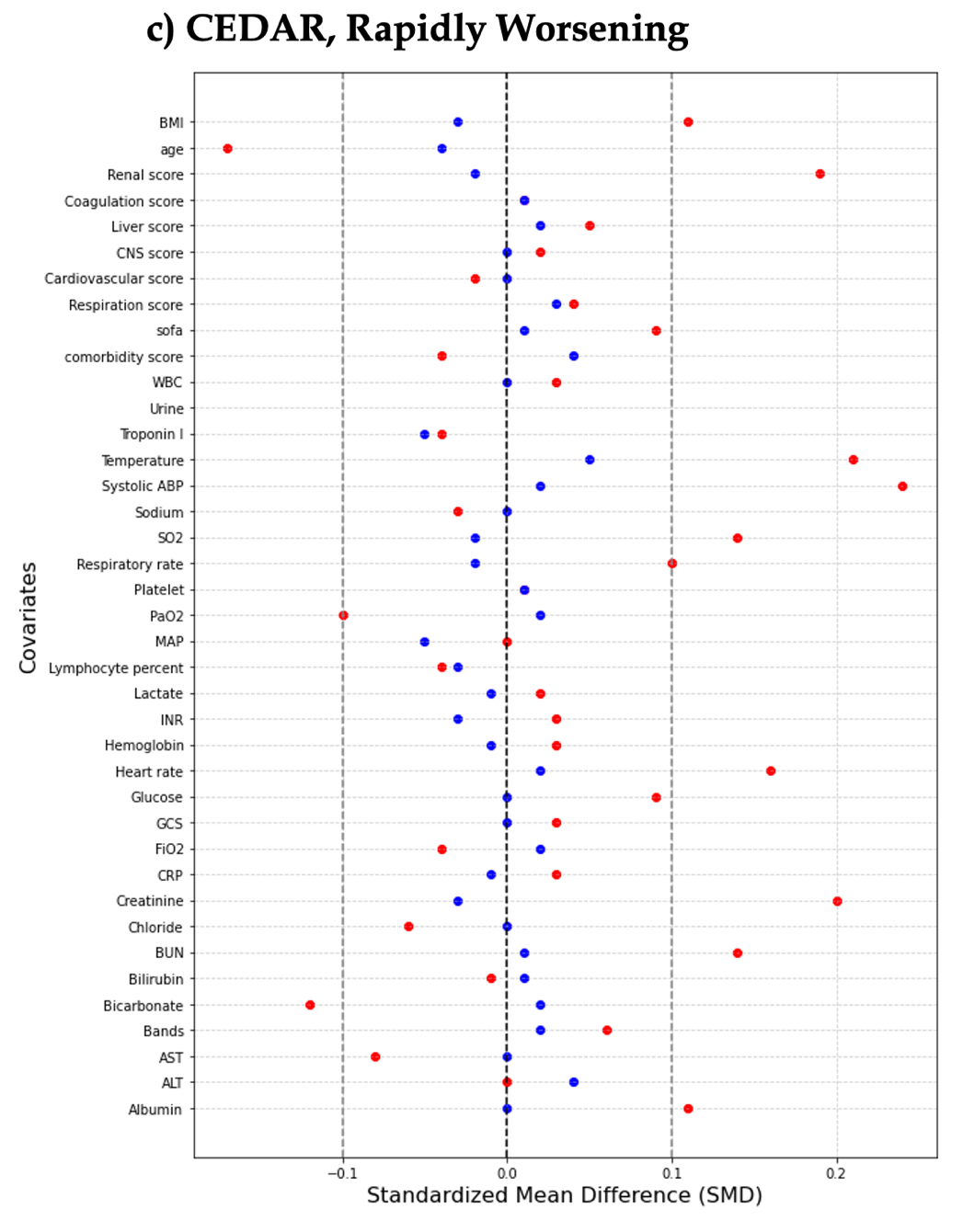

**Supplemental Figure 8**: SMD of covariates in CEDAR cohort before and after propensity score matching. SMDs are shown for 28-day mortality and time-to-discharge outcomes. Red dots indicate SMD before matching.

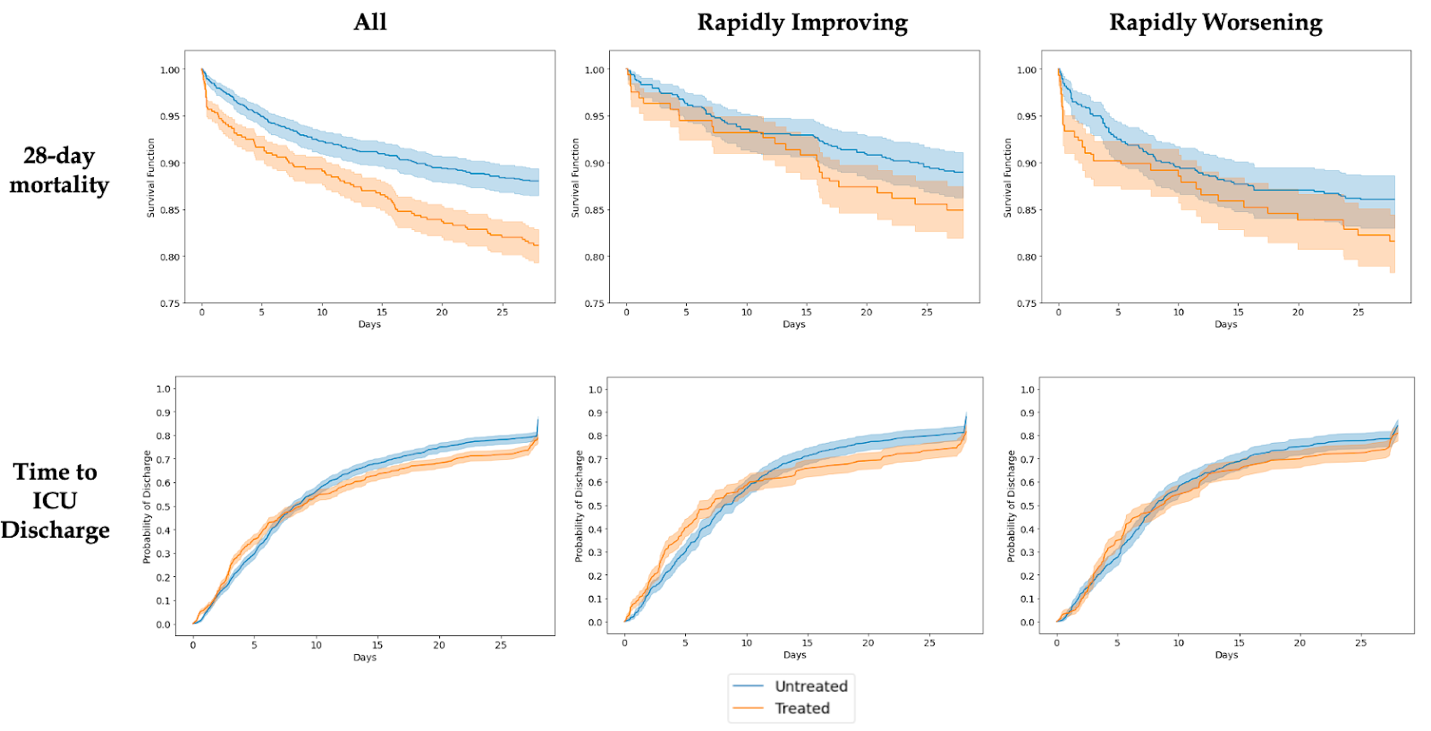

**Supplemental Figure 9.** Kaplan-Meier plots for CEDAR cohorts. Shows all, rapidly improving, and rapidly worsening groups across different outcomes

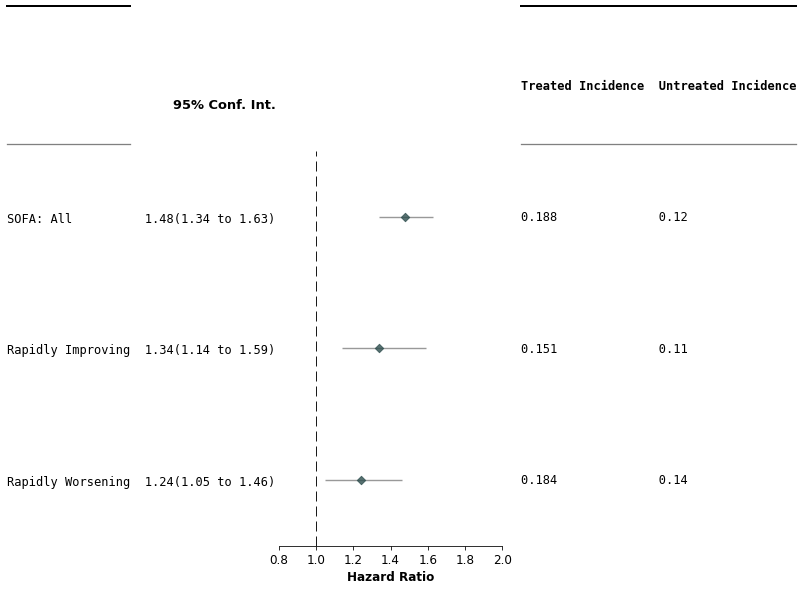

**Supplemental Figure 10. Hazard ratios and cumulative incidence for 28-day mortality outcome in CEDAR cohort.** Forest plot shows the hazard ratios and 95% confidence intervals for each patient stratification. Dashed vertical line represents a hazard ratio of 0. Cumulative incidences for treated and untreated populations are shown on the right.

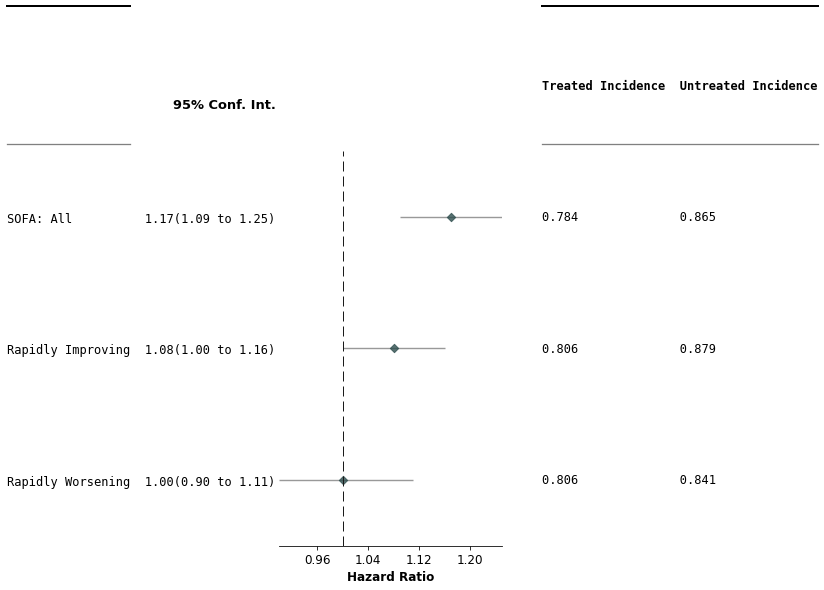

**Supplemental Figure 11. Hazard ratios and cumulative incidence for time to ICU discharge outcome in CEDAR cohort.** Forest plot shows the hazard ratios and 95% confidence intervals for each patient stratification. Dashed vertical line represents a hazard ratio of 0. Cumulative incidences for treated and untreated populations are shown on the right.

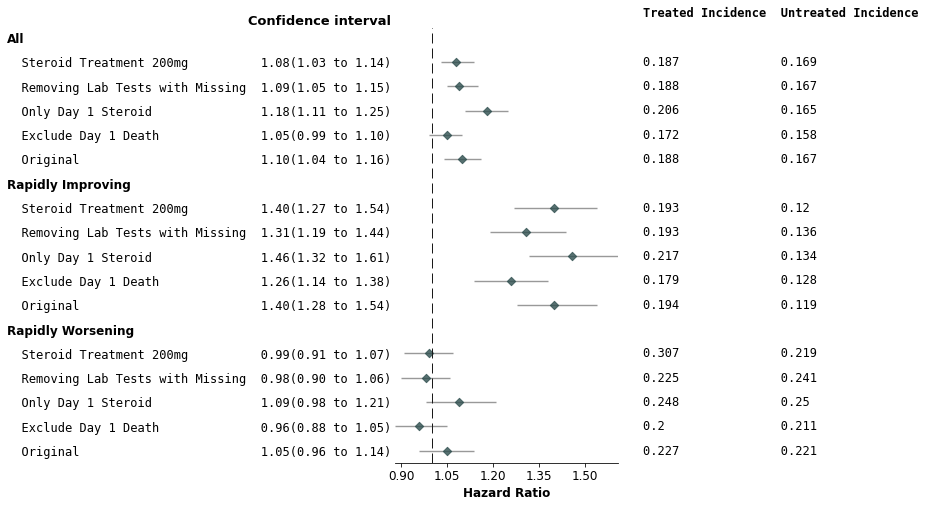

**Supplemental Figure 12. Sensitivity results for eICU-MIMIC 28-day mortality.** Forest plot shows the hazard ratios and 95% confidence intervals for each patient stratification. Dashed vertical line represents a hazard ratio of 0. Cumulative incidences for treated and untreated populations are shown on the right. The sensitivity analysis shown are (1) 200 mg instead of 160 mg treatment, (2) steroid exposure period is limited to 0 - 24hrs, (3) patients who died within Day 1 (enrollment) are removed, and (4) excluding lab test covariates with high level of missing variable

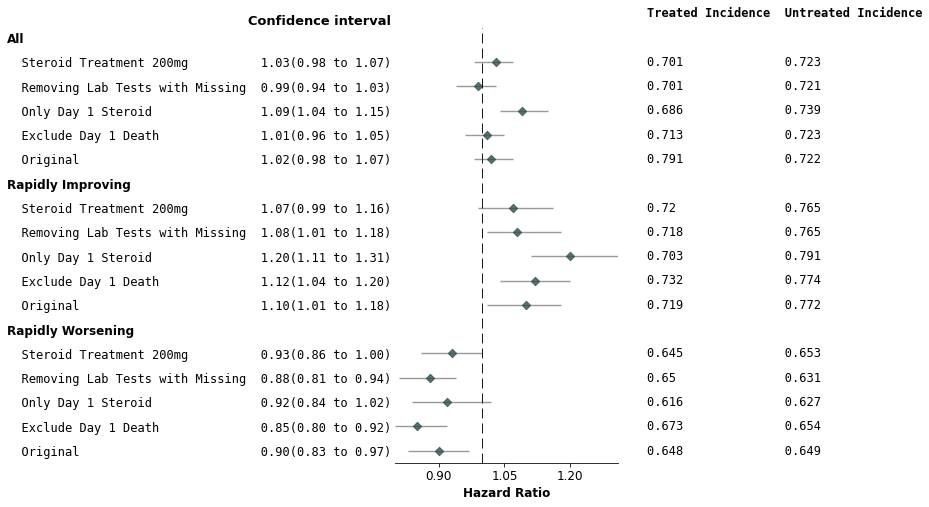

**Supplemental Figure 13. Sensitivity results for eICU-MIMIC time to ICU discharge.** Forest plot shows the hazard ratios and 95% confidence intervals for each patient stratification. Dashed vertical line represents a hazard ratio of 0. Cumulative incidences for treated and untreated populations are shown on the right. The sensitivity analysis shown are (1) 200 mg instead of 160 mg treatment, (2) steroid exposure period is limited to 0 - 24hrs, (3) patients who died within Day 1 (enrollment) are removed, and (4) excluding lab test covariates with high level of missing variable

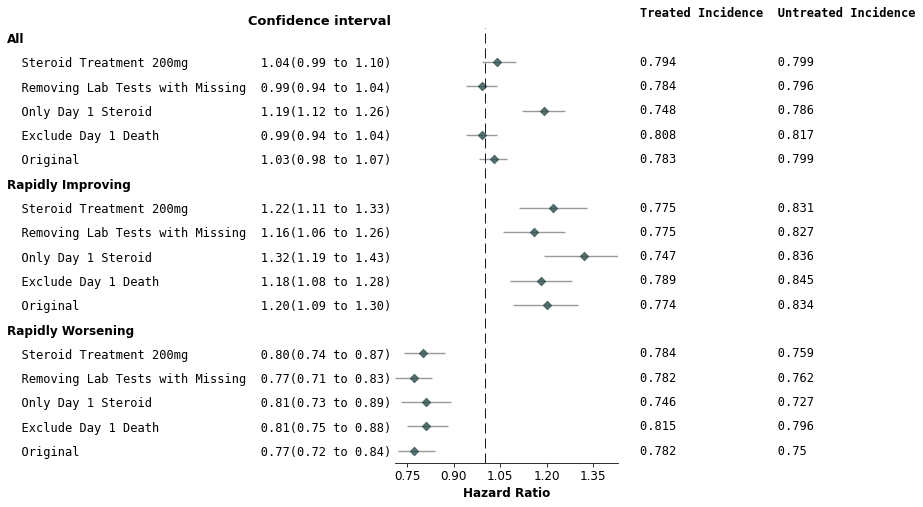

**Supplemental Figure 14. Sensitivity results for eICU-MIMIC cessation of mechanical ventilation.** Forest plot shows the hazard ratios and 95% confidence intervals for each patient stratification. Dashed vertical line represents a hazard ratio of 0. Cumulative incidences for treated and untreated populations are shown on the right. The sensitivity analysis shown are (1) 200 mg instead of 160 mg treatment, (2) steroid exposure period is limited to 0 - 24hrs, (3) patients who died within Day 1 (enrollment) are removed, and (4) excluding lab test covariates with high level of missing variable

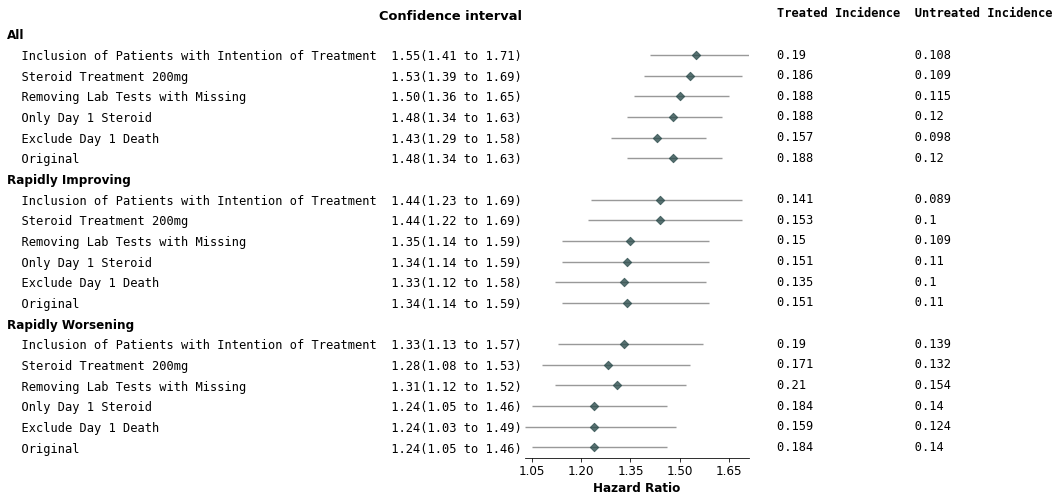

**Supplemental Figure 15. Sensitivity results for CEDAR 28-day mortality.** Forest plot shows the hazard ratios and 95% confidence intervals for each patient stratification. Dashed vertical line representing a hazard ratio of 0 is not pictured as it is lower than all lower confidence interval HRs. Cumulative incidences for treated and untreated populations are shown on the right. The sensitivity analysis shown are (1) 200 mg instead of 160 mg treatment, (2) steroid exposure period is limited to 0 - 24hrs, (3) patients who died within Day 1 (enrollment) are removed, (4) excluding lab test covariates with high level of missing variable, and (6) including patients who clinicians started treatment on Day 1 but could not complete before Day 2

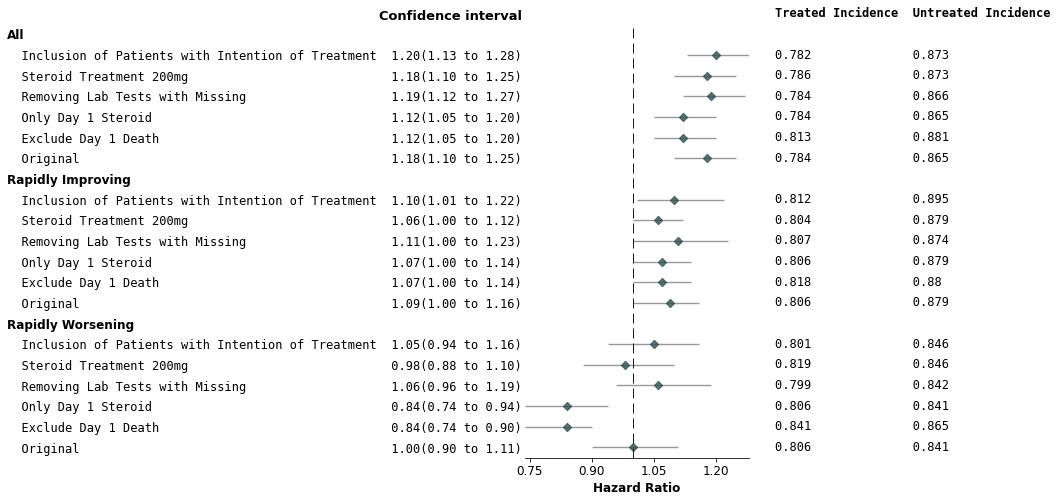

**Supplemental Figure 16. Sensitivity results for CEDAR time to ICU discharge.** Forest plot shows the hazard ratios and 95% confidence intervals for each patient stratification. Dashed vertical line represents a hazard ratio of 0. Cumulative incidences for treated and untreated populations are shown on the the right. The sensitivity analysis shown are (1) 200 mg instead of 160 mg treatment, (2) steroid exposure period is limited to 0 - 24hrs, (3) patients who died within Day 1 (enrollment) are removed, (4) excluding lab test covariates with high level of missing variable, and (6) including patients who clinicians started treatment on Day 1 but could not complete before Day 2

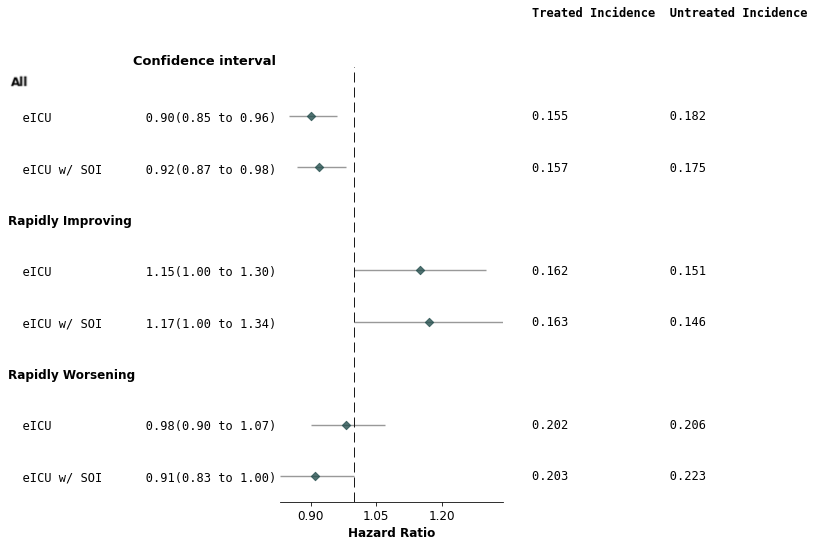

**Supplemental Figure 17. Sensitivity results for eICU 28-day mortality with and without source of infection (SOI).** Forest plot shows the hazard ratios and 95% confidence intervals for each patient stratification. Dashed vertical line represents a hazard ratio of 0. Cumulative incidences for treated and untreated populations are shown on the right. SOI stands for source of infection. Forests plot compares the HR of steroid treatment in the eICU dataset with and without the inclusion of SOI as a covariate in balancing.

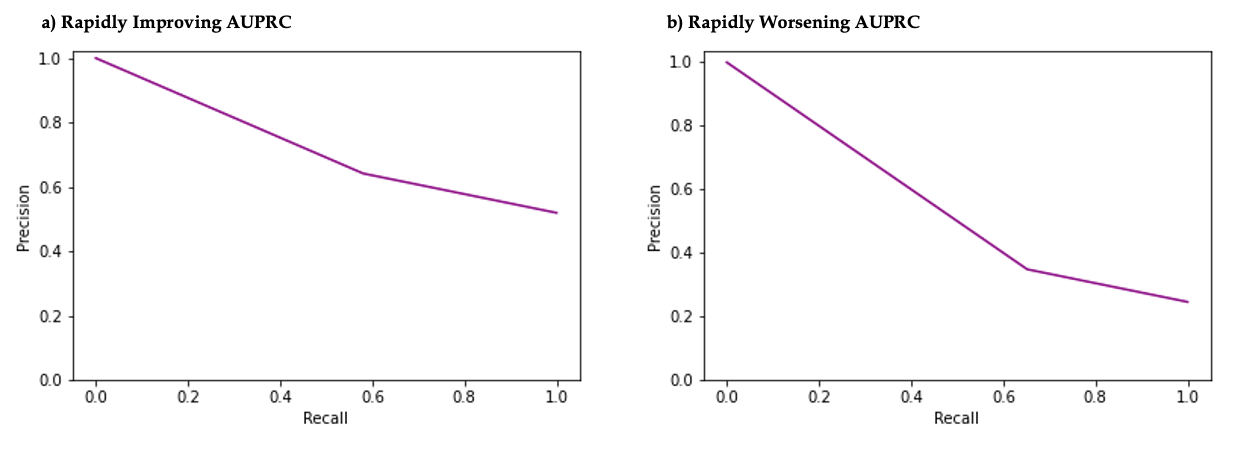

**Supplemental Figure 18**: **AUPRC curves for suphenotype logistic regression models**. (a) shows AUPRC of Rapidly Improving and (b) shown AUPRC for Rapidly Worsening.

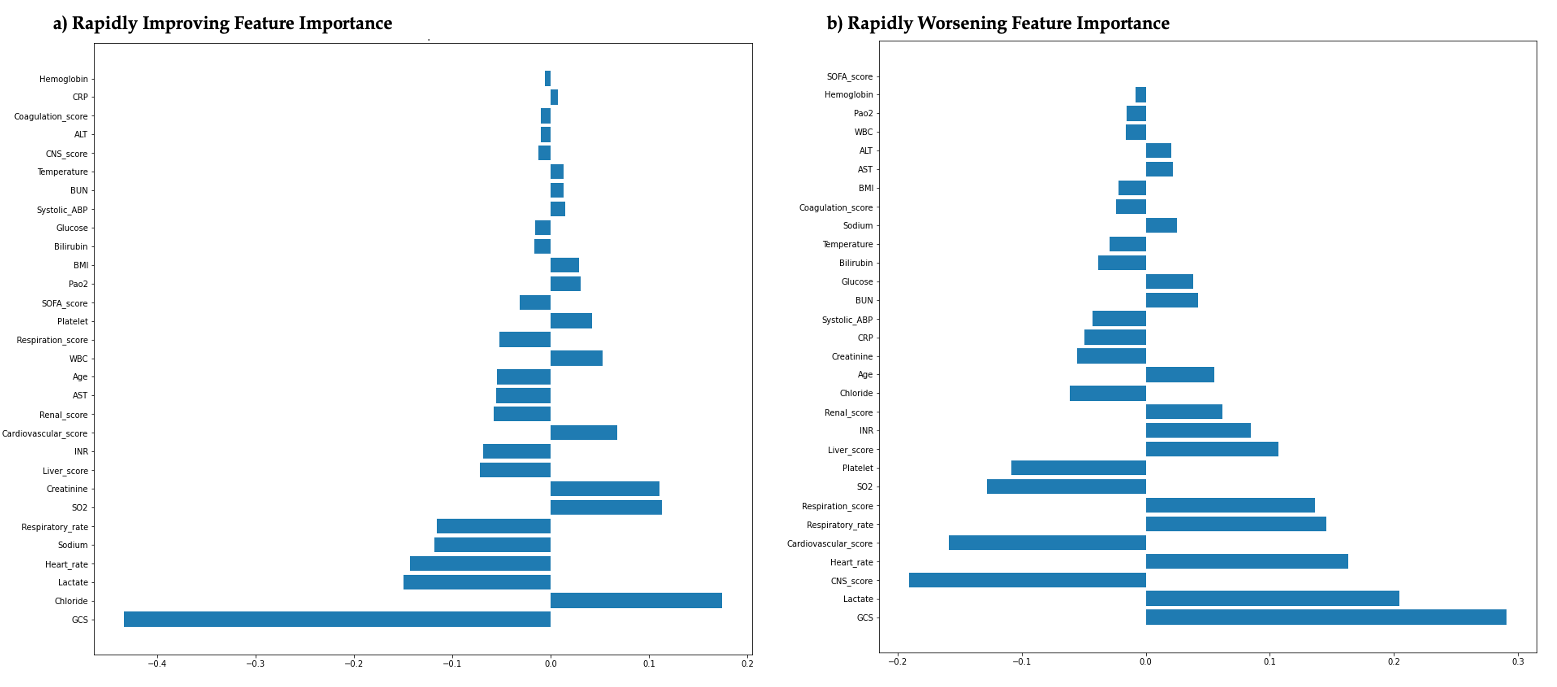

**Supplemental Figure 19**: **Feature importance plots for subphenotype logistic regression models**. (a) shows feature importance of Rapidly Improving model and (b) shown feature importance for Rapidly Worsening model.

**S3. Supplemental Text**

**Supplemental Text 1**. **Logistic Regression Model for Predicting Subphenotypes**. Features used to train subphenotype logistic regression models: SOFA_score, Respiration_score, Coagulation_score, Liver_score,  Cardiovascular_score, CNS_score, Renal_score,  CRP, Temperature, WBC, SO2, Pao2, Respiratory_rate, Heart_rate, Lactate, Systolic_ABP, BUN, Creatinine, ALT, AST, Bilirubin, GCS, Hemoglobin, INR, Platelet, Chloride, Glucose, Sodium, BMI, Age. The model for the RI group achieved an Area Under the Precision-Recall Curve (AUPRC) of 0.720, with the fraction of positives being 47.5% (as illustrated in **Supplemental Figure 18**). The model had an Area Under the Receiver Operating Curve (AUROC) of 0.615. For the RW group, the model had an AUPRC of 0.543, with the fraction of positives being 23.4% (**Supplemental Figure 18**). The model had an AUROC of 0.627. The feature importance of the two logistic regression models using the coefficients of the model was shown in **Supplemental Figure 19**. For the RI model, some important features included the Glasgow Coma Score (GCS), Chloride, Lactate, Heart rate, and Sodium levels. On the other hand, the RW model also weighted the Central Nervous System (CNS) score component of the SOFA and the Cardiovascular score of the SOFA heavily, in addition to the aforementioned features.

**Supplemental Text 2. Assessment of Unmeasured Confounding Robustness Using E-values.** Supplemental Table X presents the E-values for primary and secondary analyses across development and validation cohorts. For example, in the eICU-MIMIC cohort's rapidly improving subtype, the E-value for the 28-day mortality effect is 1.84. This indicates that an unmeasured confounder would require an association strength exceeding 1.84 with both mortality and steroid treatment to nullify the observed hazard ratio (HR) of 1.4 and alter the study's conclusions. In essence, a confounder must confer more than 1.84 times the risk to both outcomes to invalidate the calculated HR.
